# Deep learning applications for the classification of psychiatric disorders using neuroimaging data: *systematic review and meta-analysis*

**DOI:** 10.1101/2020.06.12.20129130

**Authors:** Mirjam Quaak, Laurens van de Mortel, Rajat Mani Thomas, Guido van Wingen

## Abstract

Deep learning (DL) methods have been increasingly applied to neuroimaging data to identify patients with psychiatric and neurological disorders. This review provides an overview of the different DL applications within psychiatry and compares DL model accuracy to conventional machine learning (ML). Fifty-three articles were included for qualitative analysis, primarily investigating autism spectrum disorder (ASD; n=22), schizophrenia (SZ; n=22) and attention-deficit/hyperactivity disorder (ADHD; n=9). Thirty-two of the thirty-five studies that directly compared DL to ML reported a higher accuracy for DL. Only sixteen studies could be included in a meta-regression to quantitatively compare DL and ML performance. This showed a higher odds ratio for DL models, though the comparison attained significance only for ASD. These results suggest that deep learning of neuroimaging data is a promising tool for the classification of individual psychiatric patients. The current evaluation is limited by minimal reporting of performance measures to enable quantitative comparisons, and the restriction to ADHD, SZ and ASD as current research focusses on large publicly available datasets.

## Introduction

Clinical psychiatry is based on observation and self-report which are inherently subjective. There are no biomarkers available that could enable objective diagnosis or biology-based treatment targeting. Promising approaches for the development of biomarkers include non-invasive neuroimaging techniques, such as structural or functional magnetic resonance imaging (MRI), that can capture the structure and function of the healthy and diseased brain. Over the last two decades, many neuroimaging studies have been performed to gain insight in the neural correlates of psychiatric disorders. Most of these studies have compared patients to controls and reported neuroanatomical or neurofunctional differences. This raised hopes of finding imaging biomarkers that could aid the diagnostic process. However, these studies typically relied on mass univariate analysis (group level statistical analysis) and reported group level differences in specific voxels or regions of interest (ROI) in the brain, whereas several psychiatric symptoms are best explained by network-level changes in structure and function rather than specific local alterations^1–5^.

As the vast amount of data in neuroimaging scans has made it challenging to integrate all the data available, the neuroimaging community has developed a growing interest in machine learning (ML) approaches. ML algorithms are mathematical models that are developed to learn patterns in existing data in order to make predictions on new data. A major advantage of ML techniques is their ability to take inter-regional correlations into account, enabling detection of subtle and spatially distributed effects in the brain^6^. Moreover, whereas mass-univariate results explain group differences, ML models allow statistical inference at the level of the individual and could aid individual diagnostic or prognostic decisions^7^.

Well-known pattern analysis methods, such as linear discriminant analysis (LDA), logistic regression (LR) and support vector machine (SVM) have been applied to neuroimaging data to detect psychiatric disease with varying degrees of success^7^. Classification studies using machine learning algorithms on highly dimensional neuroimaging data usually require several preprocessing steps involving feature extraction and feature selection to reduce the input dimensions^8^. These procedures require subjective feature choices that raise reproducibility issues^9^.

After breakthroughs in performance in a large variety of fields, deep learning (DL), a specific class of machine learning algorithms, has found its way into the neuroimaging community^1*^. Deep learning models are hierarchical models that achieve increasingly higher levels of abstraction and complexity by stacking consecutive nonlinear transformations (see Box 1 and Vieira et al. (2017)^10^ for an introduction) This ability makes deep learning specifically suitable for neuroimaging applications as psychiatric and neurological disorders are often characterised by complex, subtle and diffuse patterns^11^. Moreover, an essential difference between machine learning and deep learning techniques is that deep learning enables the learning of optimal feature representation from the raw data, eliminating the need for subjective feature engineering for classical machine learning techniques. This results in a more objective and less bias-prone process in deep learning ^10^.

A previous review from 2017 has shown that deep learning methods have been successfully applied in neuroimaging to classify Alzheimer, ADHD, and to predict disease conversion^10^. Since then, the advent of data-sharing initiatives and advances in deep learning have led to a large increase in deep learning applications in psychiatry. They show great promise for uncovering reproducible patterns of brain structure and function across larger and heterogeneous datasets ^12,13^.

However, also within deep learning studies, there is a large variety in techniques and processing steps that have been applied to investigate psychiatric disorders. A major challenge in the application of deep learning is the high dimensionality of neuroimaging data. Learning patterns in a high dimensional space requires thousands of labelled entries, especially for data-hungry techniques such as deep learning, whereas neuroimaging sample sizes are usually relatively small. Various studies have therefore used hand crafted input features with different levels of feature extraction along the spatial and/or temporal dimensions to reduce the input dimensionality. This diversity in approaches has made it difficult to draw any conclusions on the most optimal data input or model.

Given the high interest in DL within the field of neuroimaging for psychiatry and the wide variety of approaches, this review aims to give an overview of studies that have applied DL to neuroimaging data for the classification of psychiatric disorders. We will discuss the main themes that have emerged from our review and include a quantitative comparison of the performance of deep learning and conventional machine learning techniques. Finally, we will make a number of recommendations for future research.

## Methods

We conducted a systematic review of published studies that used deep learning approaches for diagnostic classification of psychiatric disorders using neuroimaging. The search strategy is outlined in detail in the PRISMA flow diagram in Figure 3.

**Figure 1.**
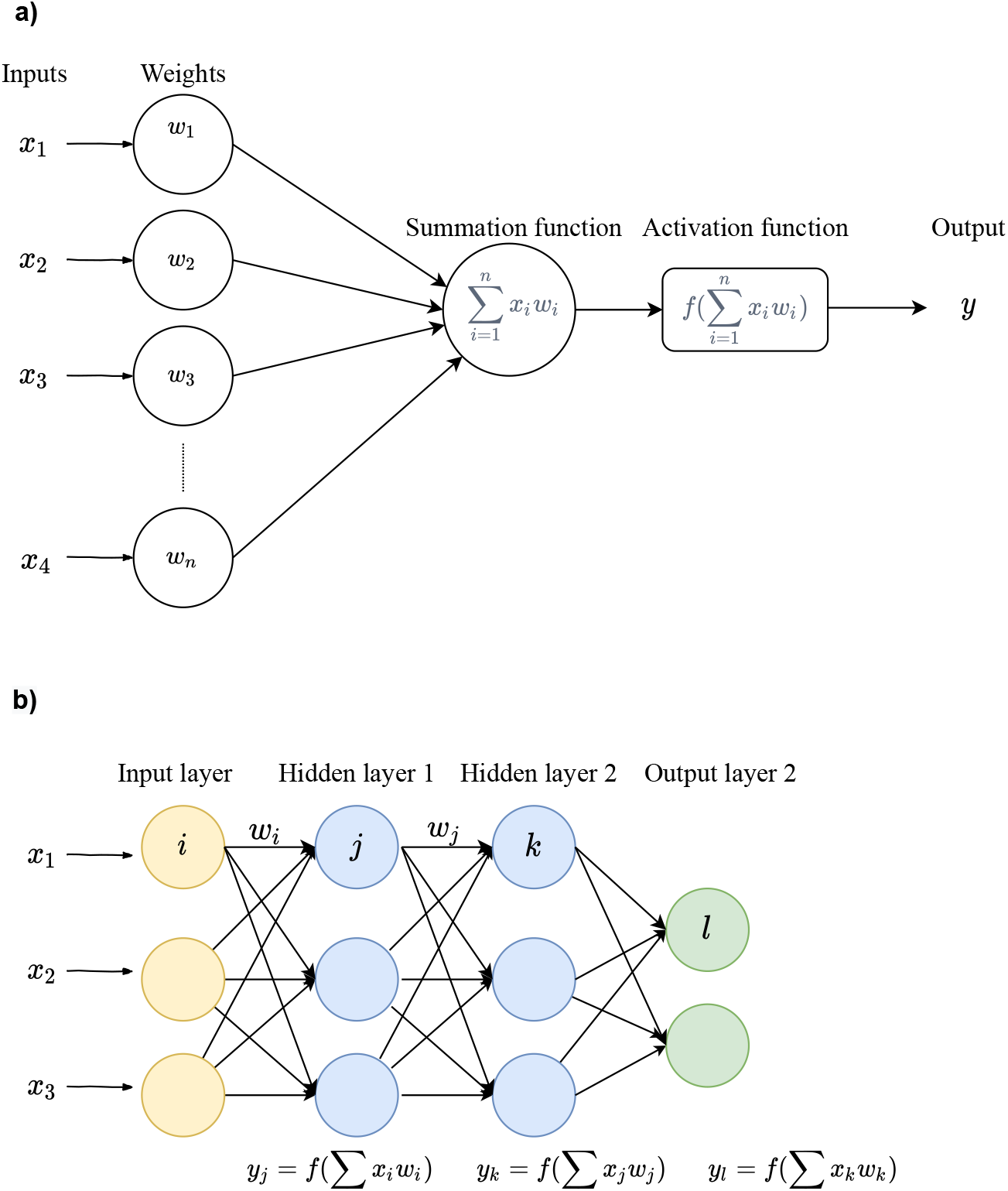
a). An artificial neuron or node. Each input *x* is associated with a weight *w*. The sum of all weighted inputs is passed onto a nonlinear activation function *f* that leads to an output *y*. b) An example of a multilayer perceptron. For each neuron in the first hidden layer, a nonlinear function is applied to the weighted sum of its inputs. The result of this transformation is the input for the consecutive layer.

**Figure 2.**
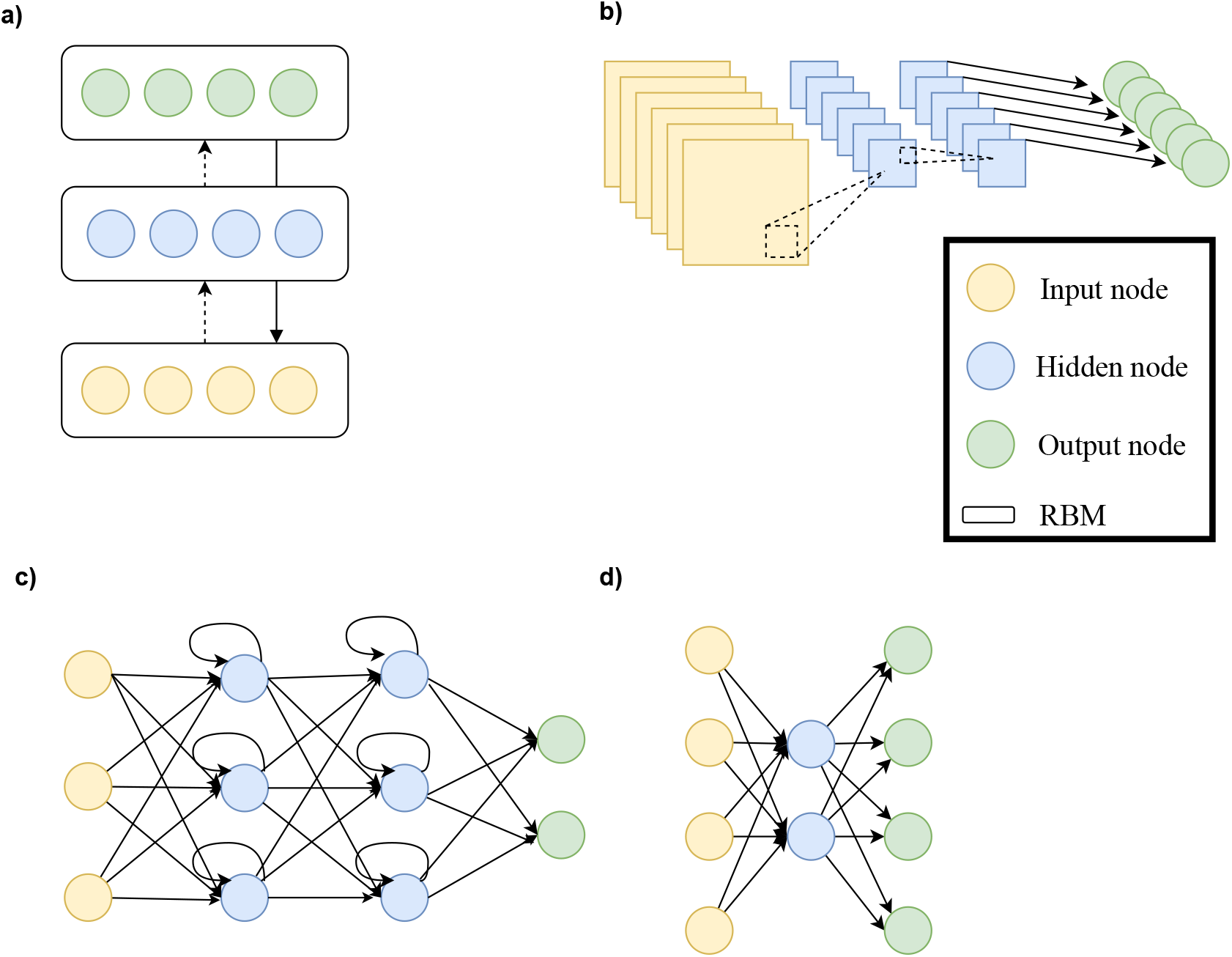
Architectural structures in deep learning.

**Figure 3.**
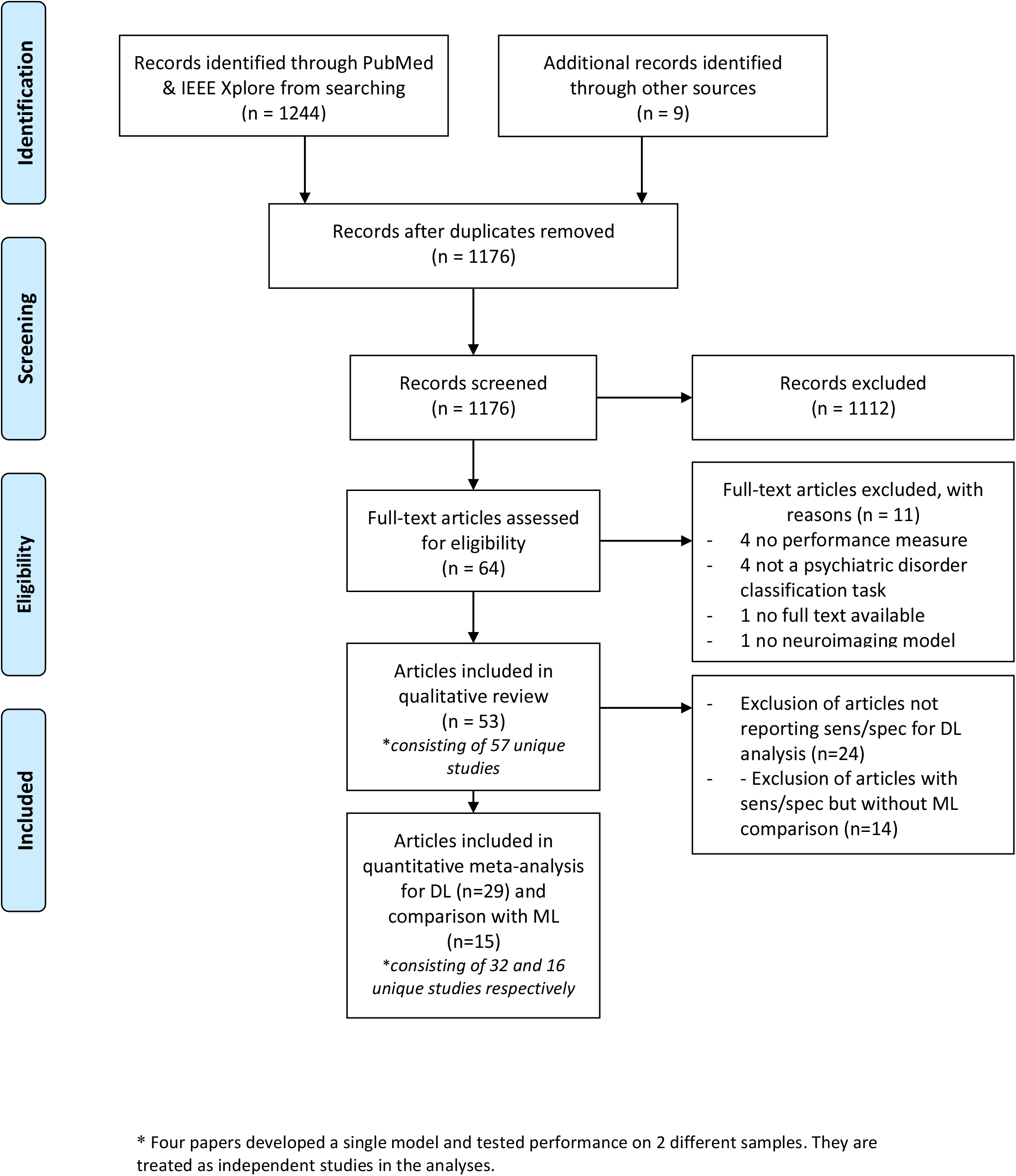
PRISMA flowchart describing the processes of literature search, study screening and selection ^87^.

### Identification

We conducted a literature search in PUBMED and IEEE Xplore using the following search string: (“deep learning” OR “deep architecture” OR “artificial neural network” OR “convolutional neural network” OR “convolutional network” OR “CNN” OR “recurrent neural network” OR “RNN” OR “Auto-Encoder” OR “Autoencoder” OR “Deep belief network” OR “DBN” OR “Restricted Boltzmann Machine” OR “RBM” OR “Long Short Term Memory” OR “Long Short-Term Memory” OR “LSTM” OR “Gated Recurrent Units” OR “GRU”) AND (psychiatry OR psychiatric OR classification OR diagnosis OR prediction OR prognosis OR outcome) AND (neuroimaging OR MRI OR “Magnetic Resonance Imaging” OR “fMRI” OR “functional Magnetic Resonance Imaging”) which is a combination of search terms used in previous reviews on deep learning in neuroimaging^10,14^. The search was limited to articles published from the 1st of January 2013 till the 30th of September 2019. In addition, articles in PubMed were identified that cited the previous systematic review on deep learning in neuroimaging of Vieira et al. (2017) ^10^. Reference lists of identified articles were further searched to select those articles that were deemed appropriate. For the scope of this study, we excluded studies using PET or EEG, although there is some evidence that DL can be used in this type of data^15^. Following this approach, 1176 studies were identified.

### Screening and Inclusion

64 Articles were eligible for full-text assessment based on title and abstract screening. Articles were included if they were a peer-reviewed full-text original research article written in English using a deep learning model for classification of a psychiatric disorder using (f)MRI. Upon full manuscript reading, 11 articles were excluded due to the lack of a clear performance measure (4), not performing a classification task of a psychiatric disorder (4), lack of a full manuscript (1), and not using a deep learning model (1), yielding a total of 53 included articles. For quantitative meta-analysis, we included 29 articles that reported sensitivity and specificity. For comparison with ML techniques, we included 15 articles that also reported sensitivity and specificity for DL and ML.

From the 53 included papers there were 4 that developed a single model and tested classification performance for 2 different samples (different psychiatric disorders) ^16–19^. These papers are included twice: they are shown independently in the two corresponding tables and are analysed as independent studies, yielding a total of 57 studies for qualitative analysis, 32 for quantitative analysis for all DL studies and 16 for quantitative meta-analysis for DL-ML comparison.

### Qualitative analysis

The included studies were grouped per disorder. We extracted data from all studies to compare a number of key aspects such as sample sizes, type of features, classifier and reported accuracies. Data extraction was done by two independent researchers and discussed if the data was inconsistent until agreement was reached. Next, we composed a narrative review of findings from included studies per disorder. Finally, we included visual summaries for all studies combined to discuss occurring themes in the literature.

### Quantitative meta-analysis

All meta-analyses were conducted using the mada and metaphor package in R. As pooling sensitivities or specificities can be misleading^20^, we have pooled studies using diagnostic odds ratios (DOR) according to the Reitsma model and the Cochrane handbook for diagnostic tests of accuracy studies^21,22^. The DOR considers both sensitivity and specificity. To visualize between study performance differences, a forest plot of the DORs is given, subdivided per disorder. In order to assess whether DL and ML models obtain different classification performances, we conducted meta-regression with classification method as covariate. We performed this meta-regression for sensitivity and false positive rates as well as DOR. In addition, the meta-regression was repeated for the largest subgroups separately.

## Results

The vast majority of studies addressed the classification of autism spectrum disorder (ASD) (n=22) or schizophrenia (SZ) (n=22). We also retrieved 9 studies for attention-deficit/hyperactivity disorder (ADHD). Finally, we included four studies about other disorders, two developed a model for major depressive disorder (MDD), one for bipolar disorder (BD) and one for conduct disorder (CD). A summary for each study including the sample size, imaging modality, DL model, and classifier performance is presented in Tables 1-4. A visual summary of reviewed studies for ASD, SZ, and ADHD is shown in Figure 4. As can be seen here, most studies (n=30) used rs-fMRI as input for their DL model. The majority of rs-fMRI studies (n=24) reduced this four-dimensional input by parcellating the brain in regions of interest (ROIs) and extracted timeseries per ROI. Most of these studies (n=16) further reduced dimensionality by analysing correlation between ROI timeseries to create a connectivity matrix (n=16). 3D input is also used often (n=12), both for fMRI, where the time dimension has been summarized, and for structural MRI. This summary also shows that a large variety of models has been deployed.

**Table 1.**
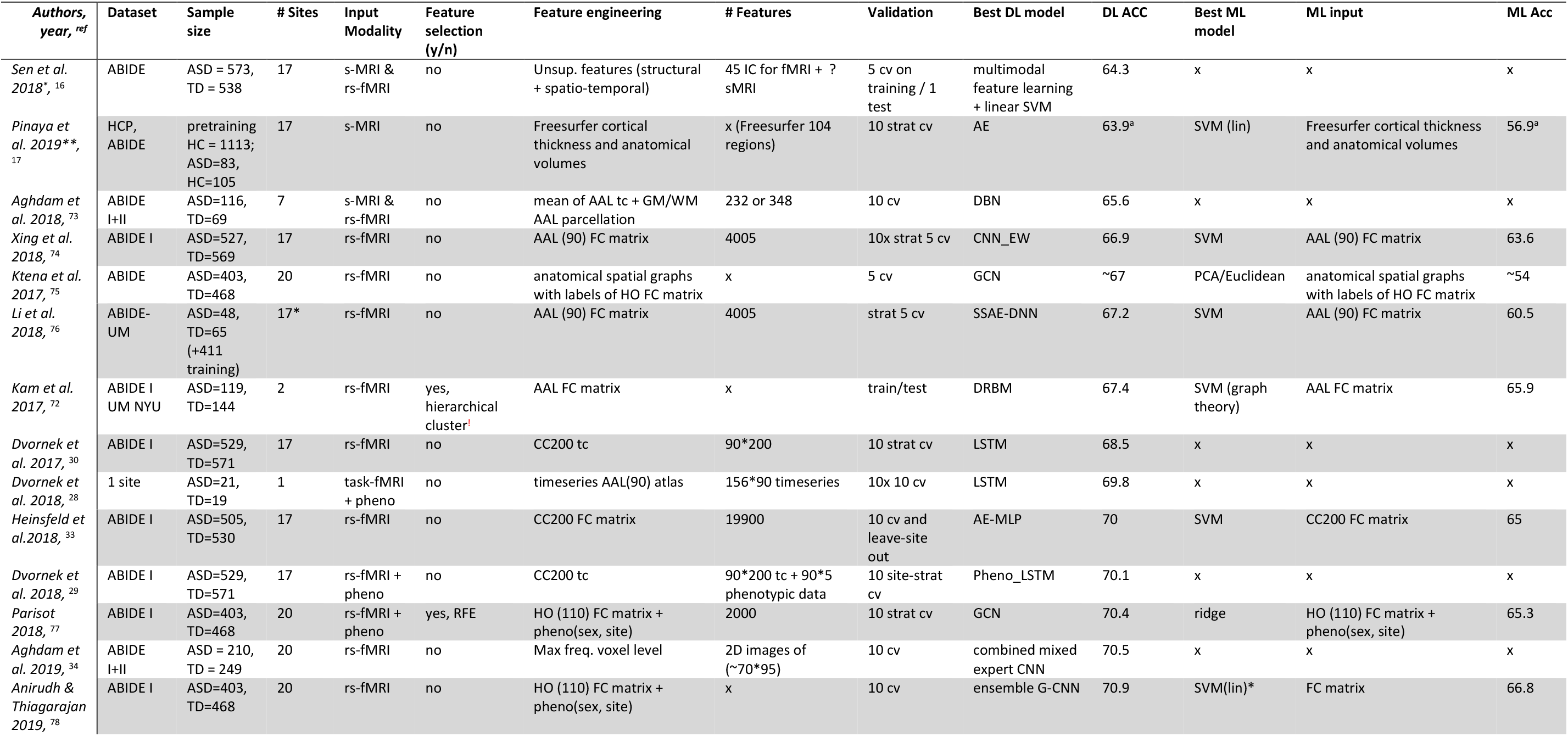
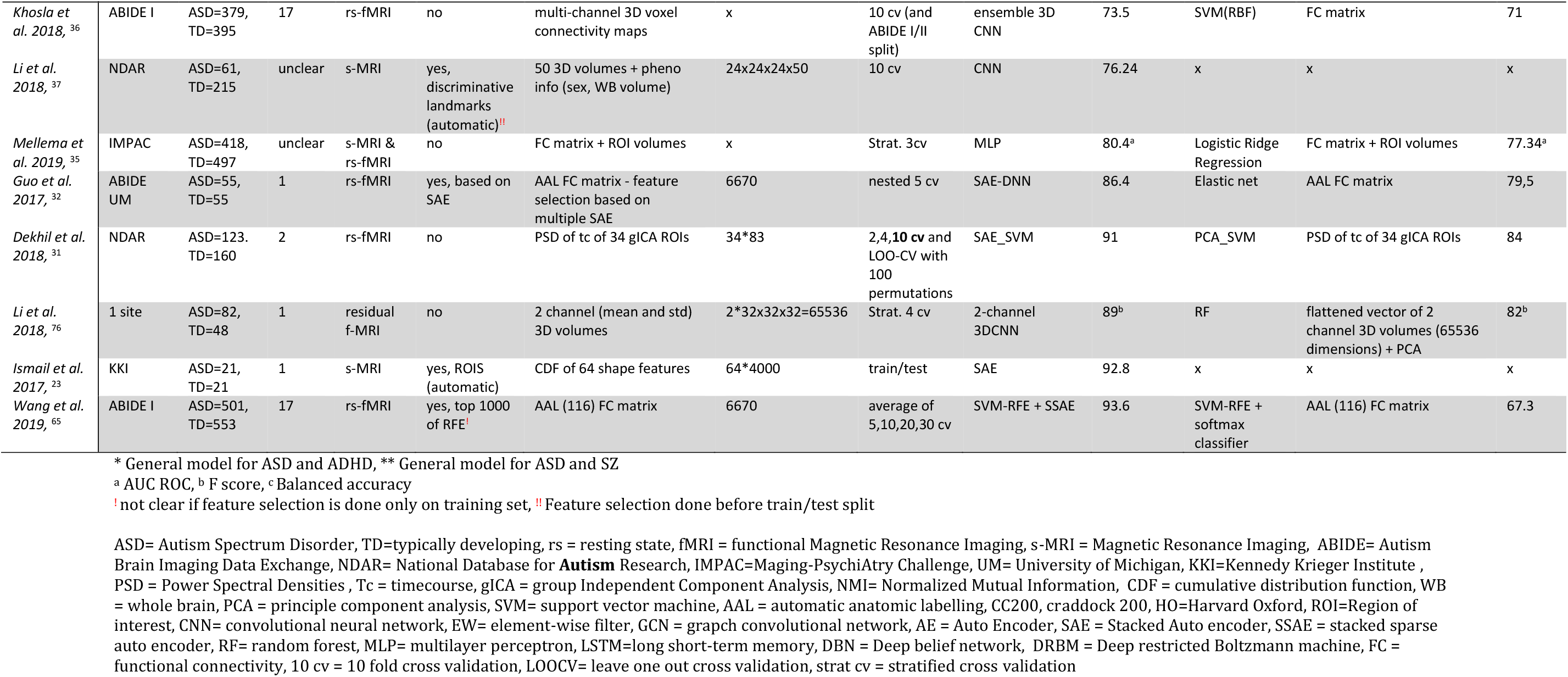
Overview of ASD studies included in this literature review

**Table 2.** Overview of SZ studies included in this literature review

| *Authors, year, <sup>ref</sup>* | Dataset | Sample size | # Sites | Input Modality | Feature selection (y/n) | Feature engineering | # Features | Validation | Best DL model | DL Acc | Best ML model | ML input | ML Acc |
| --- | --- | --- | --- | --- | --- | --- | --- | --- | --- | --- | --- | --- | --- |
| *Dakka et al. 2017, <sup>79</sup>* | 1 site | SZ=46, HC=49 | 1 | task-fMRI | no | full 4D image | x | 10 cv | LSTM | 66.4 | SVM (rbf) | 4D reduced to 1D vector | 62.1 |
| *Pinaya et al. 2019\*\*\*, <sup>17</sup>* | HCP, NUSDAST | pretraining HC = 1113 ; SZ=35,HC=40 | 1 | s-MRI | no | Freesurfer cortical thickness and anatomical volumes | x (Freesurfer 104 regions) | 10 strat cv | AE | 70.7<sup>a</sup> | SVM (lin) | Normalized Freesurfer cortical thickness and anatomical volumes | 63.7<sup>a</sup> |
| *Matsubara et al. 2019\*, <sup>18</sup>* | openfMRI | SZ=48, HC=117 | 1 | rs-fMRI | no | AAL timeseries | 116*152 | 10 cv | DGM (CVAE) | 71.3<sup>c</sup> | PCC_SCCA_SLR | AAL FC matrix | 66.4<sup>c</sup> |
| *Vyskovsky et al. 2019, <sup>57</sup>* | 1 site | SZ = 52, HC=52 | 1 | s-MRI morphometry | yes, discriminative features<sup>!</sup> | VBM and DBM Grey Matter Images | 100-10.000 | 10x LOOCV | ensemble MLP for VBM and DBM | 73.1 | SVM on VBM and DBM | VBM, DBM | 73.5 |
| *Pinaya et al. 2016, <sup>80</sup>* | 1 site | SZ = 143, HC= 83 | 1 | s-MRI |  | Freesurfer cortical thickness and anatomical volumes | x | 3 cv | DBN-DNN | 73.6<sup>c</sup> | SVM | Freesurfer cortical thickness and anatomical volumes | 68.1<sup>c</sup> |
| *Ulloa et al. 2015, <sup>81</sup>* | JHU, MPRC, IOP, WPIC | SZ = 198, HC=191 | 4 | s-MRI | no | generating sMRI images with RV generator | 55527 | 10 cv | sMRI generator + MLP | 75<sup>a</sup> | Logistic Regression | sMRI images | 70<sup>a</sup> |
| *Han 2017<sup>82</sup>* | 1 site | SZ<sup>!</sup>=39, HC=31 | 1 | rs-fMRI | no | AAL (90) FC matrix | 4005 | 10 cv | MLP | 79.3 | x | x | x |
| *Li et al. 2019, <sup>66</sup>* | 1 site | SZ=80, HC=103 | 1 | task fMRI and SNP | no | SNP loci from blood + AAL ROI | 116 | Train/test | 2 SAE + DCCA + SVM | 80.5 | x | x | x |
| *Lei et al. 2019, <sup>42</sup>* | 5 sites | SZ = 295, HC 452 | 5 | rs-fMRI | no | FC matrix 90 ROIS | 4005 | strat 5 cv | 2D CNN | 81.0<sup>c</sup> | SVM | FC matrix 90 ROIS | 81.7<sup>c</sup> |
| *Wang et al. 2019\*\*, <sup>19</sup>* | 1 site | SZ=28, HC=28 | 1 | rs- fMRI | no | based on a single 3D EPI image | 61*73*61 | 5 cv | 3D CNN | 82.2 | x | x | x |
| *Yang et al. 2019, <sup>83</sup>* | COBRE, UCLA, WUSTLE | SZ = 102, HC=120 | 3 | rs-fMRI | no | 3 ensemble inputs: sparse dictionary learning, multiple kernel mapping, AAL FC matrix | 80*20; 100*50; 116*116 | 10 cv | ensemble capsule network | 82.8 | weighted ensemble SVM | 3 ensemble inputs: sparse dictionary learning, multiple kernel mapping, AAL FC matrix | 74.2 |
| *Yan et al. 2019, <sup>39</sup>* | 7 sites | SZ = 558, HC= 542 | 7 | rs-fMRI | yes, group ICA noise<sup>!!</sup> | group ICA tc | 8500 (170 TR * 50 IC) | 10 cv and LSO | Conv + RNN | 83.2 | SVM | group ICA FC matrix (50*50) | 79.4 |

Table 2 Overview of SZ studies (Continued)
| Authors, year, <sup>ref</sup> | Dataset | Sample size | # Sites | Input Modality | Feature selection (y/n) | Feature engineering | # Features | Validation | Best DL model | DL Acc | Best ML model | ML input | ML Acc |
| --- | --- | --- | --- | --- | --- | --- | --- | --- | --- | --- | --- | --- | --- |
| Oh et al. 2019, <sup>43</sup> | 1 site | SSD = 103, HC = 41 | 1 | task-fMRI | no | 3D GLM activation map | x | 10 cv | 3D CAE-CNN | 84.4 | SVM + PCA | 3 ways: full WB, beta AAL, 40 PCA features | 70.7 |
| Yan et al. 2017, <sup>38</sup> | 7 sites | SZ = 558, HC = 542 | 7 | rs-fMRI | yes, group ICA noise<sup>!</sup> | group ICA FC matrix (50*50) | 1225 | 10 cv and LSO | DNN + LRP | 84.8 | SVMRFE | group ICA FC matrix (50*50) | 77.1 |
| Zeng et al. 2018, <sup>84</sup> | COBRE, UCLA, WUSTL, XJING1_2, AMU, Xiangya | SZ = 357, HC= 377 | 7 | 6 rs-fMRI, 1 task fMRI | no | FC of diff atlases (ROI: 176, 160, 116) |  | 10 cv + leave site out validation | DANS with 3 atlas features fusion at label level | 85.0 | RFE-LDA | selected features from correlation matrices 3 atlases label level fusion; | 80.9 |
| Kim et al. 2016, <sup>85</sup> | COBRE | SZ= 50, HC=50 | 1 | rs-fMRI | no | group ICA FC matrix (116*116) | 6670 | 10 x nested 5 cv | 2 SAE + DNN | 86.5 | SVM (lin) | FC matrix GICA | 76.9 |
| Plis et al. 2014, <sup>11</sup> | JHU, MPRC, IOP, WPIC | SZ = 198, HC=191 | 4 | s-MRI | no | RBM feature learning | 60645 voxel GM images | 10 cv | RBM of 3 layers + Logistic regression for classification | 91<sup>b</sup> | x | x | x |
| Chyzhyk 2015, <sup>46</sup> | COBRE | SZ=72, HC=74 | 1 | rs-fMRI | Yes, evolutionary selection algorithm | VHMC map | 86559 | 10 cv | Ensemble of ELM | 91.2 | RF on ReHo | ReHO selected C map | 80.9 |
| Patel 2016, <sup>45</sup> | COBRE | SZ=72, HC=74 | 1 | rs-fMRI | yes, filter out inactive or noisy GM voxels | AAL (116) timeseries |  | 10 cv | SAE_SVM | 92 | x | x | x |
| Srinivasagopalan 2019, <sup>44</sup> | Kaggle dataset | SZ = 69, HC=75 | 1 | s-MRI & rs-fMRI | yes, ICA noise selection | FC maps ICA brain maps derived from GM concentration | 411 | Train/test | MLP | 94.4 | RF | 55 selected features with RFE and RF | 83.3 |
| Qureshi et al. 2019, <sup>40</sup> | COBRE | SZ = 72, HC=72 | 1 | rs-fMRI | yes, group ICA noise<sup>!</sup> | 3D-ICA | 15 | 10 cv | 3DCNN | 98.0 | x | x | x |
| Qureshi et al. 2017, <sup>41</sup> | COBRE | SZ = 72, HC=72 | 1 | s-MRI & rs-fMRI | yes, group ICA noise<sup>!</sup> | structural ROI, global functional connectivity, group ICA, kernel PCA with spatial ICA maps | 748 | nested 10 by 10 cv | ELM | 99.3 | SVM-L | structural ROI, global functional connectivity, group ICA, kernel PCA with spatial ICA maps | 77.8 |

\* General model SZ and BD, \*\* General model SZ and ADHD, \*\*\* General model SZ and ASD
SZ<sup>1</sup> early onset Schizophrenia
<sup>a</sup> AUC ROC, <sup>b</sup> F score, <sup>c</sup> Balanced accuracy
<sup>!</sup> not clear if feature selection is done only on training set, <sup>!!</sup> Feature selection done before train/test split

**Table 3.**
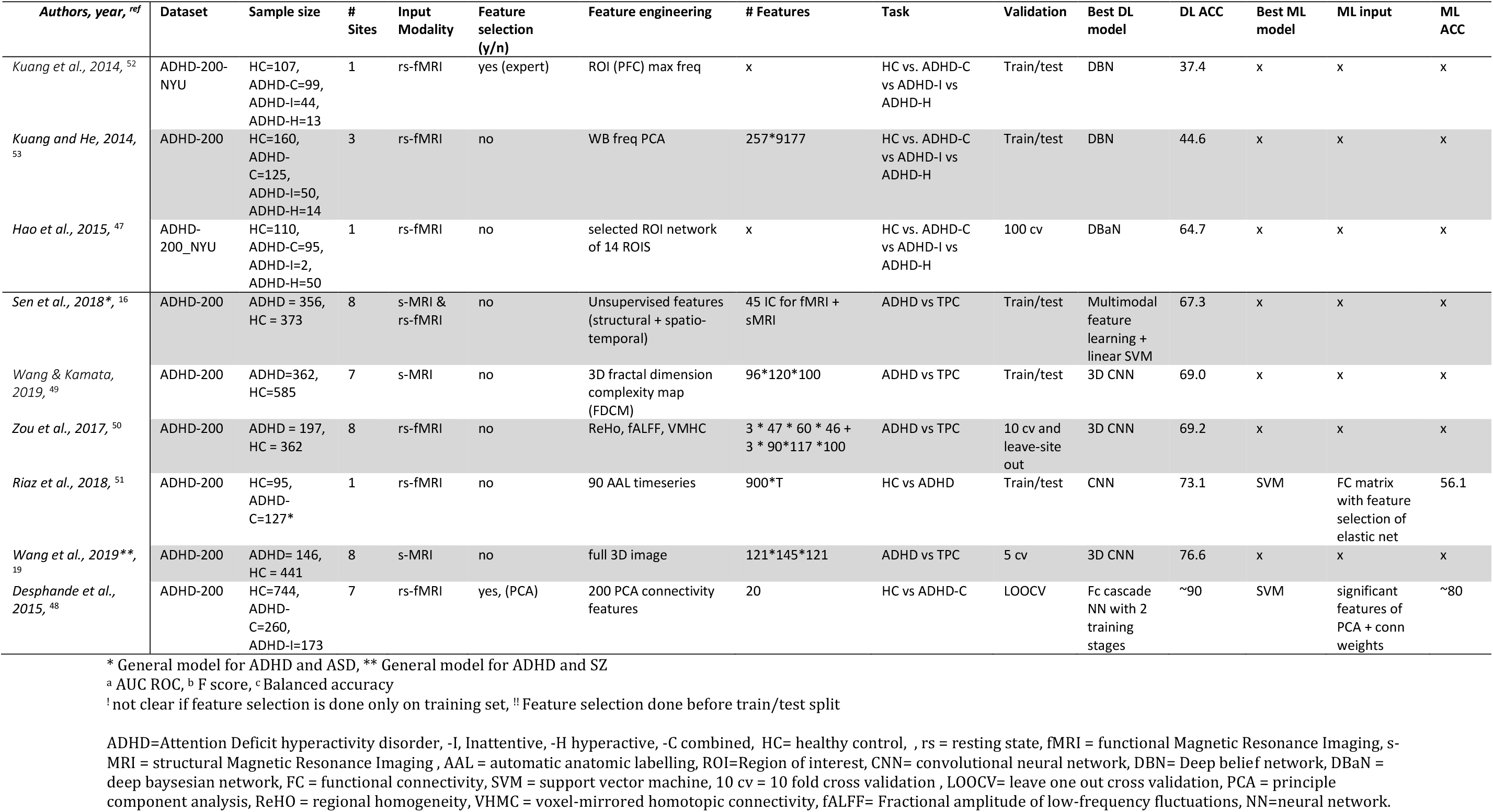
Overview of ADHD studies included in this literature review

| *Authors, year, <sup>ref</sup>* | Dataset | Sample size | # Sites | Input Modality | Feature selection (y/n) | Feature engineering | # Features | Task | Validation | Best DL model | DL ACC | Best ML model | ML input | ML ACC |
| --- | --- | --- | --- | --- | --- | --- | --- | --- | --- | --- | --- | --- | --- | --- |
| *Kuang et al., 2014, <sup>52</sup>* | ADHD-200-NYU | HC=107, ADHD-C=99, ADHD-I=44, ADHD-H=13 | 1 | rs-fMRI | yes (expert) | ROI (PFC) max freq | x | HC vs. ADHD-C vs ADHD-I vs ADHD-H | Train/test | DBN | 37.4 | x | x | x |
| *Kuang and He, 2014, <sup>53</sup>* | ADHD-200 | HC=160, ADHD-C=125, ADHD-I=50, ADHD-H=14 | 3 | rs-fMRI | no | WB freq PCA | 257*9177 | HC vs. ADHD-C vs ADHD-I vs ADHD-H | Train/test | DBN | 44.6 | x | x | x |
| *Hao et al., 2015, <sup>47</sup>* | ADHD-200_NYU | HC=110, ADHD-C=95, ADHD-I=2, ADHD-H=50 | 1 | rs-fMRI | no | selected ROI network of 14 ROIS | x | HC vs. ADHD-C vs ADHD-I vs ADHD-H | 100 cv | DBaN | 64.7 | x | x | x |
| *Sen et al., 2018\*, <sup>16</sup>* | ADHD-200 | ADHD = 356, HC = 373 | 8 | s-MRI & rs-fMRI | no | Unsupervised features (structural + spatio-temporal) | 45 IC for fMRI + sMRI | ADHD vs TPC | Train/test | Multimodal feature learning + linear SVM | 67.3 | x | x | x |
| *Wang & Kamata, 2019, <sup>49</sup>* | ADHD-200 | ADHD=362, HC=585 | 7 | s-MRI | no | 3D fractal dimension complexity map (FDCM) | 96*120*100 | ADHD vs TPC | Train/test | 3D CNN | 69.0 | x | x | x |
| *Zou et al., 2017, <sup>50</sup>* | ADHD-200 | ADHD = 197, HC = 362 | 8 | rs-fMRI | no | ReHo, fALFF, VMHC | 3 * 47 * 60 * 46 + 3 * 90*117 *100 | ADHD vs TPC | 10 cv and leave-site out | 3D CNN | 69.2 | x | x | x |
| *Riaz et al., 2018, <sup>51</sup>* | ADHD-200 | HC=95, ADHD-C=127* | 1 | rs-fMRI | no | 90 AAL timeseries | 900*T | HC vs ADHD | Train/test | CNN | 73.1 | SVM | FC matrix with feature selection of elastic net | 56.1 |
| *Wang et al., 2019\*\*, <sup>19</sup>* | ADHD-200 | ADHD= 146, HC = 441 | 8 | s-MRI | no | full 3D image | 121*145*121 | ADHD vs TPC | 5 cv | 3D CNN | 76.6 | x | x | x |
| *Desphande et al., 2015, <sup>48</sup>* | ADHD-200 | HC=744, ADHD-C=260, ADHD-I=173 | 7 | rs-fMRI | yes, (PCA) | 200 PCA connectivity features | 20 | HC vs ADHD-C | LOOCV | Fc cascade NN with 2 training stages | ~90 | SVM | significant features of PCA + conn weights | ~80 |

\* General model for ADHD and ASD, \*\* General model for ADHD and SZ
<sup>a</sup> AUC ROC, <sup>b</sup> F score, <sup>c</sup> Balanced accuracy
<sup>!</sup> not clear if feature selection is done only on training set, <sup>!!</sup> Feature selection done before train/test split
ADHD=Attention Deficit hyperactivity disorder, -I, Inattentive, -H hyperactive, -C combined, HC= healthy control, , rs = resting state, fMRI = functional Magnetic Resonance Imaging, s-MRI = structural Magnetic Resonance Imaging, AAL = automatic anatomic labelling, ROI=Region of interest, CNN= convolutional neural network, DBN= Deep belief network, DBaN = deep bayesian network, FC = functional connectivity, SVM = support vector machine, 10 cv = 10 fold cross validation, LOOCV= leave one out cross validation, PCA = principle component analysis, ReHo = regional homogeneity, VMHC = voxel-mirrored homotopic connectivity, fALFF= Fractional amplitude of low-frequency fluctuations, NN=neural network.

**Table 4.** Overview of BD, CD, MDD studies included in this literature review

| Authors,<br>year,<sup>ref</sup> | Disorder | Dataset | Sample size | #<br>Sites | Input<br>Modality | Feature<br>selection<br>(y/n) | Feature<br>engineering | # Features | Task | Validation | Best DL model | DL Acc | Best ML model | ML input | ML<br>Acc |
| --- | --- | --- | --- | --- | --- | --- | --- | --- | --- | --- | --- | --- | --- | --- | --- |
| Matsubara<br>et al.,<br>2019*,<sup>18</sup> | Bipolar<br>Disorder<br>(BD) | openfMRI | BD=46,<br>HC=117 | 1 | rs-fMRI | no | AAL timeseries | 116*152 | BD vs<br>HC | 10 cv | DGM (CVAE) | 64.0<sup>c</sup> | PCC_Kendall_LLE_Cmeans | AAL FC matrix | 62.2<sup>c</sup> |
| Zhang et<br>al. 2019,<sup>54</sup> | Conduct<br>disorder<br>(CD) | 1 site | CD = 60,<br>HC=60 | 1 | s-MRI | no | full 3D image<br>with<br>augmentation | 121*145*121 | CD VS<br>HC | 5 cv | 3D CNN | 85 | SVM(lin) | VBM | 77 |
| Pominova<br>et al.,<br>2018,<sup>55</sup> | Major<br>Depressive<br>Disorder<br>(MDD) | 1 site | MDD = 25,<br>HC=25 | 1 | rs-fMRI | yes,<br>cleaned<br>data<br>(unclear) | full 4D image | 52*62*52*133 | MDD vs<br>HC | 5 cv | 3DConvLSTM | 73 | x | x | x |
| Miholca &<br>Onicas,<br>2017,<sup>56</sup> | Major<br>Depressive<br>Disorder<br>(MDD) | openfMRI | MDD=19,<br>HC=20 | 1 | task-fMRI | yes, task<br>related<br>ROI<sup>!</sup> | task-related<br>param. of<br>selected ROIs | x | MDD vs<br>HC | LOOCV | MLP | 92.3 | RAR based classifier | task-related<br>param. of<br>selected ROIs | 94.8 |

\* General model for BD and SZ
<sup>a</sup> AUC ROC, <sup>b</sup> F score, <sup>c</sup> Balanced accuracy
<sup>!</sup> not clear if feature selection is done only on training set, <sup>!!</sup> Feature selection done before train/test split
BD = Bipolar Disorder, CD = Conduct Disorder, MDD = Major Depressive Disorder, HC= healthy control, , rs = resting state, fMRI = functional Magnetic Resonance Imaging, s-MRI = Magnetic Resonance Imaging, AAL = automatic anatomic labelling, ROI=Region of interest, DGM = Deep neural generative model, CVAE = conditional variational auto encoder, CNN= convolutional neural network, ConvLSTM = convolutional Long Short-Term Memory, MLP = multilayer perceptron, FC = functional connectivity, SVM = support vector machine, RAR = Relational association rules, VBM = Voxel based morphometry, LLE = locally linear embedding, 10 cv = 10 fold cross validation, LOOCV= leave one out cross validation

**Figure 4.**
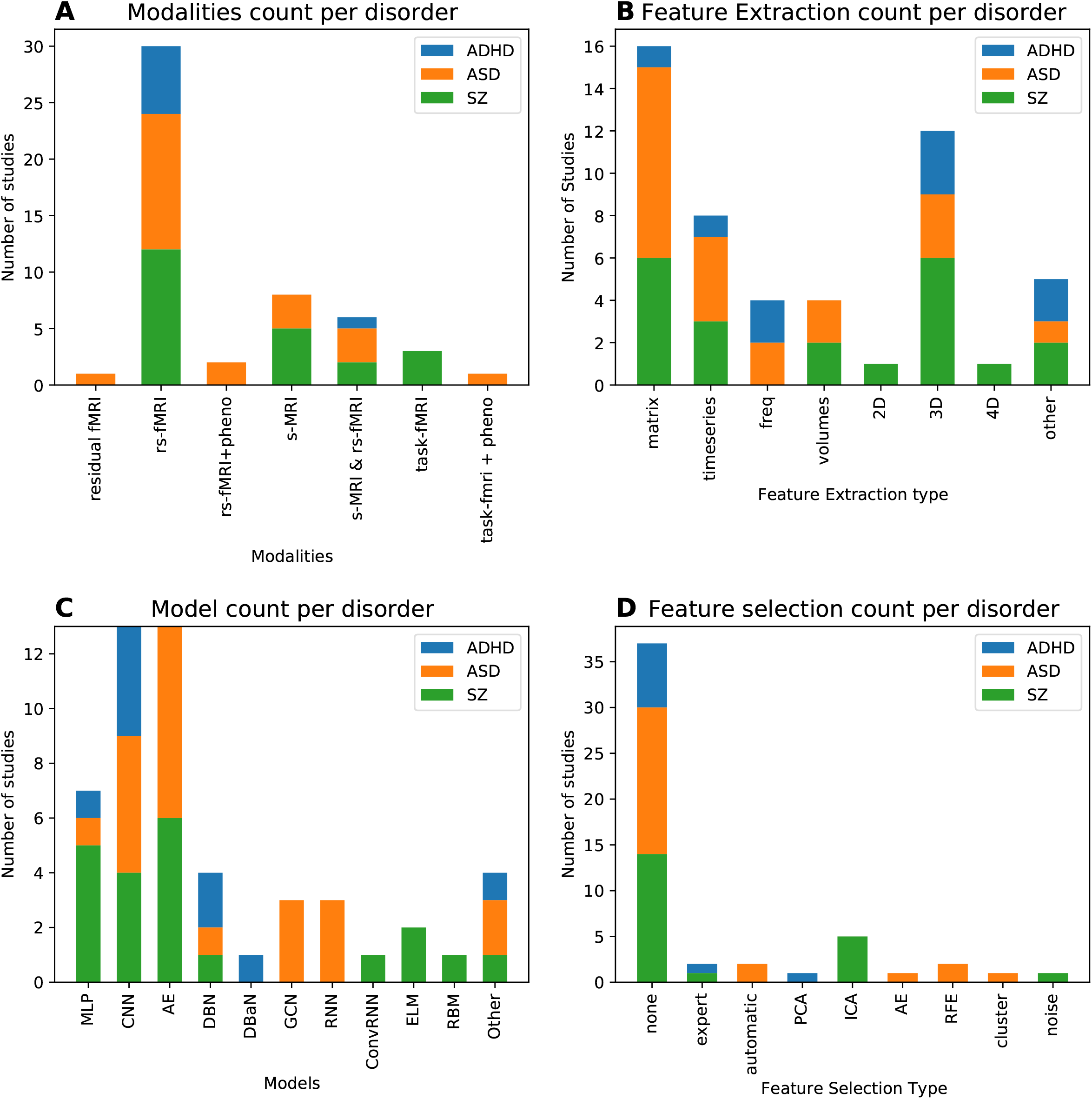
Visual summary of articles reviewed grouped by the three most investigated disorders ADHD, ASD and SZ. A) Number of articles on different modalities; B) Number of articles of different feature extraction, C) number of articles on different DL models, D) Number of articles on different feature selection procedures.

### Autism spectrum disorder (ASD)

Twenty-three studies have applied DL for classification of ASD with accuracies ranging from 50-94. As shown in Table 1, eighteen studies have used data from the Autism Brain Imaging Data Exchange (ABIDE), a data-sharing initiative involving >20 different scanning sites. The ABIDE features over 2000 structural and functional MRI scans of autistic and typically developing children and came out in two releases: ABIDE-I and II. Even though these studies have used the same dataset, there is a large difference in subsets used, with sample sizes ranging from 110 to 1054. As shown in Figure 4, three studies have used only structural MRI (s-MRI) as input. The study with the highest reported accuracy using s-MRI of Ismail et al. (2017)^23^ used a stacked auto encoder (SAE) on cumulative distribution function (CDF) of shape features and reached up to 92.8% accuracy. However, this is also the study with the smallest study size (n=42) and they did not report cross-validated results. Of the studies using fMRI, the vast majority used resting state (rs-) fMRI. When controlling for the physiological and motion from task f-MRI and using residual fMRI, Li et al. (2018)^24^ obtained 89% accuracy. They used a 3D convolutional model on 3D brain volumes where the time dimension is summarized in mean and standard deviation of voxel’s timecourses per time window. This study also reports the highest accuracy for studies without doing any feature selection. As can be seen in Table 1, when accuracy performances are getting higher (lower in the Table), feature selection is done more often. One needs to be careful with concluding that feature selection is beneficial for performance, as several studies have done feature selection on the whole sample instead of properly selecting features only on the training set. One of them reported a very high accuracy of 93.6 on the full ABIDE I dataset, using a stacked sparse autoencoder on selected features of a functional connectivity (FC) matrix. They applied SVM-RFE for the selection of 1000 features. However, this appears to be done on the entire dataset without keeping the test set separately. This increases the risk of overfitting, complicates model interpretation, and may produce optimistic results ^25–27^.

When looking at input features, the FC matrix is often used as input; a total of 10 ASD studies have applied a DL model on FC matrices of different atlases, most commonly the AAL, Craddock or HO atlas. This is probably because the ABIDE provides extracted timecourses for these atlas parcellations. Instead of focusing on FC matrices, four studies have incorporated the time dimension and worked on timeseries as input data^28–31^. In three different studies by Dvornek et al. (2017, 2018, 2018) ^28–30^, they have experimented with the optimal input for Long Short-Term Memory (LSTM) models. They have shown that incorporating phenotypic data such as sex, IQ and age improves performance by 3 per cent ^29^. The highest performance on timeseries input is reported by a study from Dekhil et al. (2018)^31^. They transformed timeseries into power spectral densities (PSD) for 34 group independent component analysis (ICA) spatial maps and used sparse auto encoders (SAE) to reduce the input dimensionality so it could be fed into an SVM. They obtained a high accuracy of 88%, but on a relatively homogeneous dataset with 2 different scanning sites (as compared to >20 in ABIDE).

Two other studies applied auto-encoders (AE) to reduce the dimensionality of the data in an unsupervised way; Guo et al. (2017)^32^ applied a stacked AE to the FC matrix and obtained an accuracy of 86.36 on one site (UM) of the ABIDE. Instead of reducing feature dimensions, Heinsfeld et al. (2018)^33^ initialized an MLP with AE weights using FC matrices and obtained an accuracy of 70 on the entire ABIDE I dataset (n=1035).

There are three studies that have incorporated both structural and functional MRI as input to the DL model (Sen et al. (2018)^16^, Aghdam et al. (2018)^34^, Mellema et al. (2019)^35^). Of these three Mellema et al. (2019)^35^ reported the highest accuracy of 80.4 on a large dataset (n=915) by inputting FC and ROI volume values into an MLP. A major part of their success seems to be due to their multimodal input, as even a simplistic logistic regression obtained an accuracy of 77.3. Finally, there are three studies that have worked on 3D input data (Khosla et al. (2018)^36^, Li et al. (2018)^37^, Li et al. (2018)^24^). Khosla et al. (2018)^36^ used the largest, most heterogeneous dataset (n=774, sites=17) and achieved 73.5 by using an ensemble of 3D Convolutional Neural Networks. Overall, a wide variety of input, models and subset of the data has been used, making it difficult to make direct comparisons between studies.

### Schizophrenia (SZ)

Similar to the other disorders, the first papers on deep learning for schizophrenia classification appeared in 2016 and in the last 3 years many papers have followed. We included twenty-two studies for SZ classification with an accuracy range of 66-99 that are shown in Table 2. In contrast to ASD, there is a large variety in datasets used despite different data sharing efforts such as the MCIC and COBRE. Most sample sizes are relatively small as compared to the ABIDE or ADHD-200. The largest studies of Yan et al. (2017, 2019)^38,39^ with a cohort of 1100 subjects report accuracies over 80%, which is relatively high as compared to the classification performances on the full ABIDE dataset. Yet, the SZ samples might be more homogeneous as it only consists of seven different scanning sites. Yan et al. (2017, 2019)^38,39^ have reported a model on FC matrices of group ICA spatial maps as well its timeseries. Their first model on the FC matrices using an MLP with layer wise relevance propagation (LRP) outperformed the Convolutional Recurrent Network on timeseries, but the difference is small: 84.8% vs 83.2%. Both studies do group ICA on the whole sample and filter out noise before splitting the data into training and test sets. The test set is thereby also used for feature engineering, hereby making the model ‘peek’ into test data and making it more susceptible to overfitting,^27^ though the influence of including test data on the group ICs may be minimal.

The group ICA feature selection has been done by more studies for SZ classification. In two studies of Qureshi it et al. (2017, 2019)^40,41^, it is also not explicitly mentioned whether this is done on the training set only. They report the highest SZ classification accuracy of 99.3% on the COBRE dataset using an Extreme Learning Machine (ELM) on a multimodal input of structural MRI and rs-fMRI^41^. Their performance on using rs-fMRI only of 98.1% is only slightly lower^40^. Here, they applied 3D convolutions to 3D ICA volumes. There are two other studies applying a convolutional network, both reporting accuracies over 80%^42,43^.

One other study used a multimodal input from structural and resting-state functional MRI and achieved an accuracy of 94.4 with a normal MLP of 3 layers on FC and ICA maps ^44^. Seven studies have used the COBRE dataset, of which the highest accuracies reported are from Qureshi et al. (2017, 2019)^40,41^, followed by Patel et al. ^45^ with an accuracy of 92%. They trained an SAE on each ROI timeseries to obtain an encoded vector that could be fed into an SVM. Chyzyk et al. (2015)^46^ obtained a similarly high accuracy of 91% with a very different approach; using an ensemble of ELMs on 3D voxel-mirrored homotopic connectivity (VHMC) maps that passed an evolutionary algorithm for feature selection. Remarkably, one study on restricted boltzmann machine (RBM) of Plis et al. (2016) also obtained an F score of 0.91 using only structural MRI on a larger dataset (n=389).

One study applied transfer learning; Pinaya et al. (2019)^17^ trained deep auto encoders on data from the human connectome project (HCP) to create a normative model of structural MRI. They then used the normative model to estimate neuroanatomical deviations in individual patients in SZ as well as ASD. Using these deviations for classification, they obtained an accuracy of 70.7% for SZ and 63.9% for ASD.

### ADHD

We included nine studies on ADHD classification. As shown in Table 3, they all have used the ADHD-200 dataset. Nevertheless, sample size varies and ranges from 349-1167 subjects. Three studies have performed classification of the ADHD subtypes (inattentive, hyperactive or both) with accuracies ranging from 27 to 65 (chance level of 25% for classification of 4 different groups). The highest performance for subtype classification is reported by Hao et al. (2016)^47^ that achieved 64.7% on a constructed Bayesian network on the max frequencies ROIS from rs-fMRI data. For bivariate classification of ADHD the highest accuracy is reported by Deshpande et al. (2015)^48^. They used a fully connected cascade neural network on PCA connectivity features and obtained around 90% accuracy. Their PCA connectivity selection seemed to work well as they obtained a relatively high accuracy of 80% using SVM on this data, which was higher than all the other reported accuracies of DL models.

Using a convolutional neural network on structural MRI, Wang et al. (2019)^19^ applied 3D convolutions and obtained an accuracy of 77.6%. They also tested their model on SZ data and obtained an accuracy of 82.2% for SZ. One other study by Sen et al. (2018)^16^ developed a general model for psychiatric disorder classification and tested it on ADHD and ASD and obtained 68% and 63% respectively.

Three other studies deployed convolutions for classification^49,50^, all with different inputs: AAL timeseries^51^, a combination of ReHo, fALFF and VHMC ^50^ or 3D structural maps ^49^. There does not appear to be a large difference between using rs-fMRI or structural MRI in these studies, but they are difficult to compare as they have used different subsets of the ADHD-200 and applied different validation procedures.

Remarkably, four out of nine studies do not perform cross-validation but train their model once on training data and then report the performance on test data^49,51–53^. This might be because the ADHD-200 dataset started off as a competition and provides this train/test split.

### Other disorders

We included four studies that investigated classification of other disorders, which are summarized in Table 4. These four studies have relatively small sample sizes, ranging from 49 to 163. One study of Matsubara et al. (2015)^18^ developed a general model for classification of fMRI and tested this for both schizophrenia and bipolar disorder (BD). They used the AAL timeseries and obtained a balanced accuracy of 64% for BD (and 71.3 for SZ). Zhang et al. (2019)^54^ applied 3D convolutions on structural MRI to classify conduct disorder (CD) with an accuracy of 85%. Two studies classified major depressive disorder (MDD)^55,56^. Miholca & Onicas obtained an accuracy of 92% using an MLP on task fMRI, but they selected features on the whole dataset, including test data. Pominova et al. (2018)^55^ is one of the rare studies that did not perform feature engineering, but applied a 3DConvLSTM model on full 4D fMRI data. They obtained an accuracy of 73% on a relatively small dataset of 50 subjects.

### Effect of sample size and number of sites

The effect of sample size on accuracy is illustrated in Figure 5. Although there is no obvious linear relation, there is a significant negative monotonic relation between sample size and accuracy when combining all the studies (*r*_*s*_*=*-0.32, *p*=0.02). Though when splitting the data per disorder, these trends did not reach significance and were even absent or in opposite direction (ASD: *r*_*s*_*=*-0.42, *p*=0.05; SZ: *r*_*s*_*=*0.02, *p*=0.94; ADHD: *r*_*s*_*=*0.43, *p*=0.24). When splitting the data for number of sites, no significant relation was observed (see Fig S1, S2).

**Figure 5.**
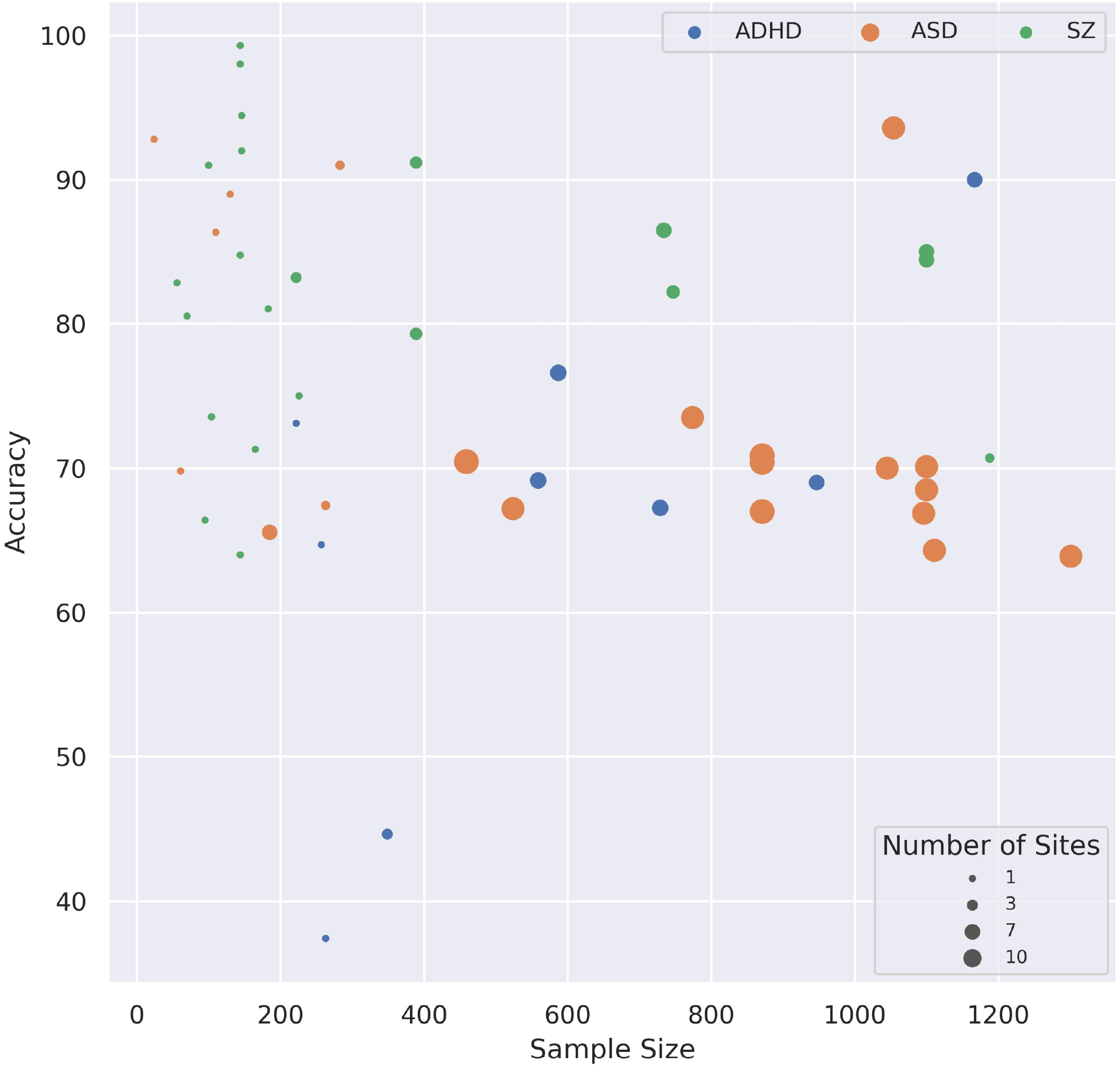
Figure 5 Scatterplot of accuracy for different sample sizes, the size of the dots indicates the number of scanning sites included in the sample.

Naturally, larger samples usually involve more scanning sites, thus more heterogeneity in the data. It also shows that SZ studies have more studies with high performances (>90% accuracy), but that most of these are conducted on small datasets. ASD studies often involve large sample sizes with many scanning sites, which could be explained by the publicly available ABIDE dataset.

### Deep learning vs. machine learning

A total of thirty-five studies included in this review compared a DL model against a conventional machine learning method (such as SVM, LR or RF). The results of these studies are shown in Figure 6. For thirty-two of the thirty-five included studies (91%), DL showed improved performance as compared to ML. Given the heterogeneity of the input of the models, it is difficult to identify specific characteristics of the studies associated with greater improvement when applying DL. The difference seems to go up whenever DL models are gaining higher performances. Only three studies report lower performance for DL than ML^42,56,57^: Lei et al. (2019)^42^ compared many different models of which SVM achieved the highest performance on the AAL FC matrix. The 2D convolutional neural network only performed slightly worse (difference of 0.7%). In Vyskovsky et al. (2019)^57^ an ensemble of MLPs was outperformed by an ensemble of SVMs for first episode schizophrenia classification with a marginal difference of 0.4%. Finally, in Miholca & Onicas (2017)^56^ a new kind of ML technique using relational association rules achieved a 2.6% better accuracy score than an MLP.

**Figure 6.**
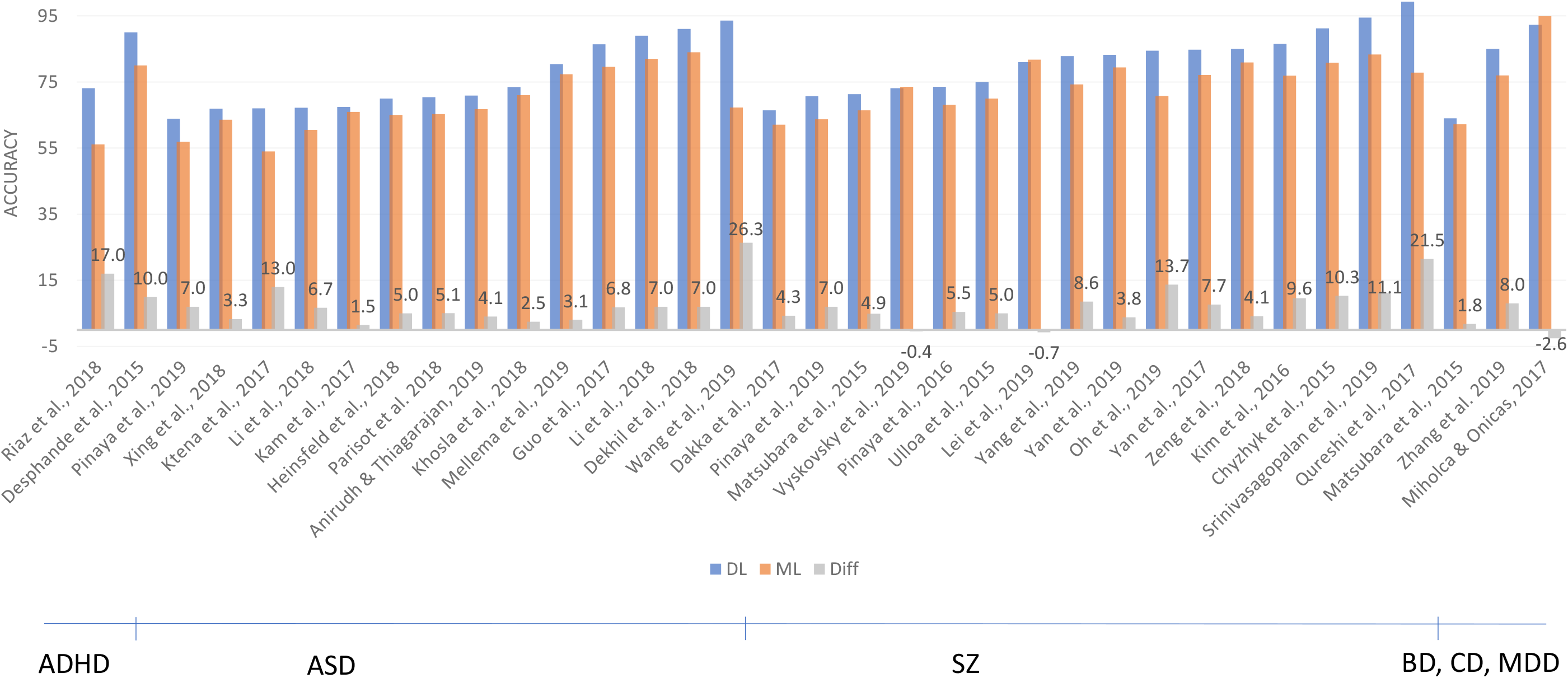
Results of studies comparing DL and conventional ML models. The graph shows the accuracies (or other reported performance scores: AUC, balanced Acc, F score) for DL models in blue and ML models in orange. The difference between the two groups is depicted in grey.

### Quantitative meta-analysis

To test whether DL techniques achieved significantly higher performances than ML techniques, we performed a quantitative meta-analysis for 16 studies that 1) directly compared a DL model with ML and 2) reported sensitivity and specificity results to perform meta-analysis for bivariate classification. Figure 7 shows an illustrative forest plot of the included studies. The pooled DOR for deep learning models was 2.51 [2.04, 2.97] versus 2.04 [1.59, 2.50] for machine learning models. To test whether this difference was significant we performed a random-effect meta-regression for type of model, for which the results are presented in Table 5. Although DL had a higher odds ratio, the difference between the two estimates was not significant (*p=0*.*166*). When comparing sensitivity and false positive rates (fpr) separately according to the Reitsma model, DL had a higher sensitivity, but the difference was again non-significant (*p=0*.*151*). The false positive rates were higher for machine learning models (*p=0*.*013*), but this did not remain significant after Bonferroni correction for multiple comparisons.

**Table 5.**
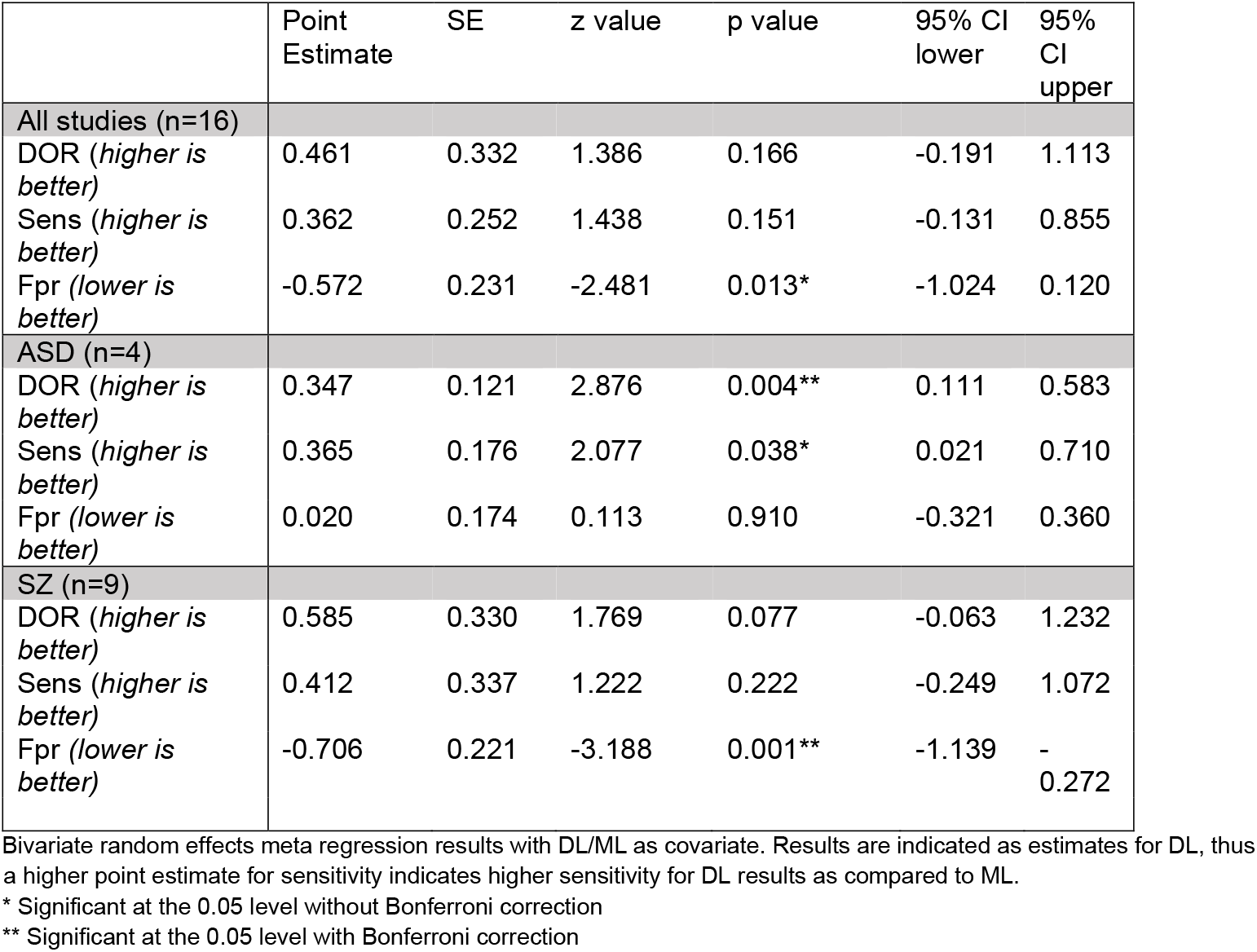
bivariate random-effect meta-regression with DL/ML as covariate

|  | Point Estimate | SE | z value | p value | 95% CI lower | 95% CI upper |
| --- | --- | --- | --- | --- | --- | --- |
| **All studies (n=16)** |  |  |  |  |  |  |
| DOR (*higher is better*) | 0.461 | 0.332 | 1.386 | 0.166 | -0.191 | 1.113 |
| Sens (*higher is better*) | 0.362 | 0.252 | 1.438 | 0.151 | -0.131 | 0.855 |
| Fpr (*lower is better*) | -0.572 | 0.231 | -2.481 | 0.013* | -1.024 | 0.120 |
| **ASD (n=4)** |  |  |  |  |  |  |
| DOR (*higher is better*) | 0.347 | 0.121 | 2.876 | 0.004** | 0.111 | 0.583 |
| Sens (*higher is better*) | 0.365 | 0.176 | 2.077 | 0.038* | 0.021 | 0.710 |
| Fpr (*lower is better*) | 0.020 | 0.174 | 0.113 | 0.910 | -0.321 | 0.360 |
| **SZ (n=9)** |  |  |  |  |  |  |
| DOR (*higher is better*) | 0.585 | 0.330 | 1.769 | 0.077 | -0.063 | 1.232 |
| Sens (*higher is better*) | 0.412 | 0.337 | 1.222 | 0.222 | -0.249 | 1.072 |
| Fpr (*lower is better*) | -0.706 | 0.221 | -3.188 | 0.001** | -1.139 | -0.272 |

Bivariate random effects meta regression results with DL/ML as covariate. Results are indicated as estimates for DL, thus a higher point estimate for sensitivity indicates higher sensitivity for DL results as compared to ML.
\* Significant at the 0.05 level without Bonferroni correction
\*\* Significant at the 0.05 level with Bonferroni correction

**Figure 7.**
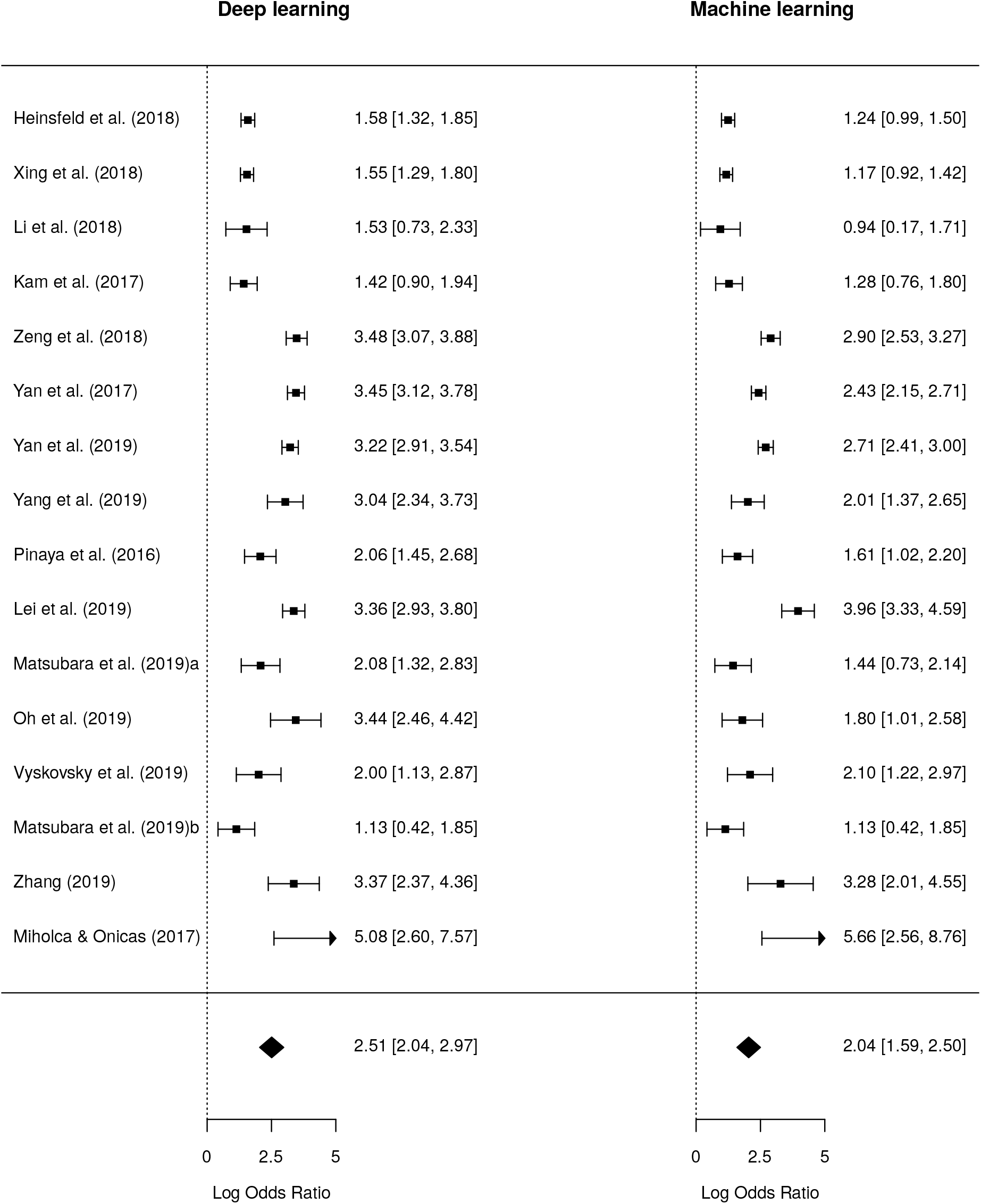
Forest plot of diagnostic odds ratio for deep learning and machine learning comparison.

When pooling studies that investigated the same disorder, there was a significant increase in DL performance in ASD (n=4) as measured by increased odds ratio (*p=0*.*004*). A similar effect was present for sensitivity (*p=0*.*038*), which did not remain significant after Bonferroni correction. For SZ (n=9), there was only a significant difference for false positive rate (*p=0*.*001*) with ML results showing higher fpr.

#### Pooled DOR per disorder

The univariate forest plot of DOR of all studies included in the meta-analysis is shown in Figure 8. The total pooled DOR of DL studies was 2.76 [95% CI= 2.24-3.27]. Pooled DOR for ADHD studies was lowest with 1.60 [95% CI=0.73-2.60], followed by ASD with a pooled DOR of 2.15 [95% CI=1.22-3.08] and the highest for SZ studies with a pooled DOR of 3.38 [95% CI=2.82-3.95]. Again, it can be seen that there is large variety in performance of models within a disorder, which is probably caused by sample variance as inter-study differences are present in population, modalities, type of DL model, feature selection and engineering technique.

**Figure 8.**
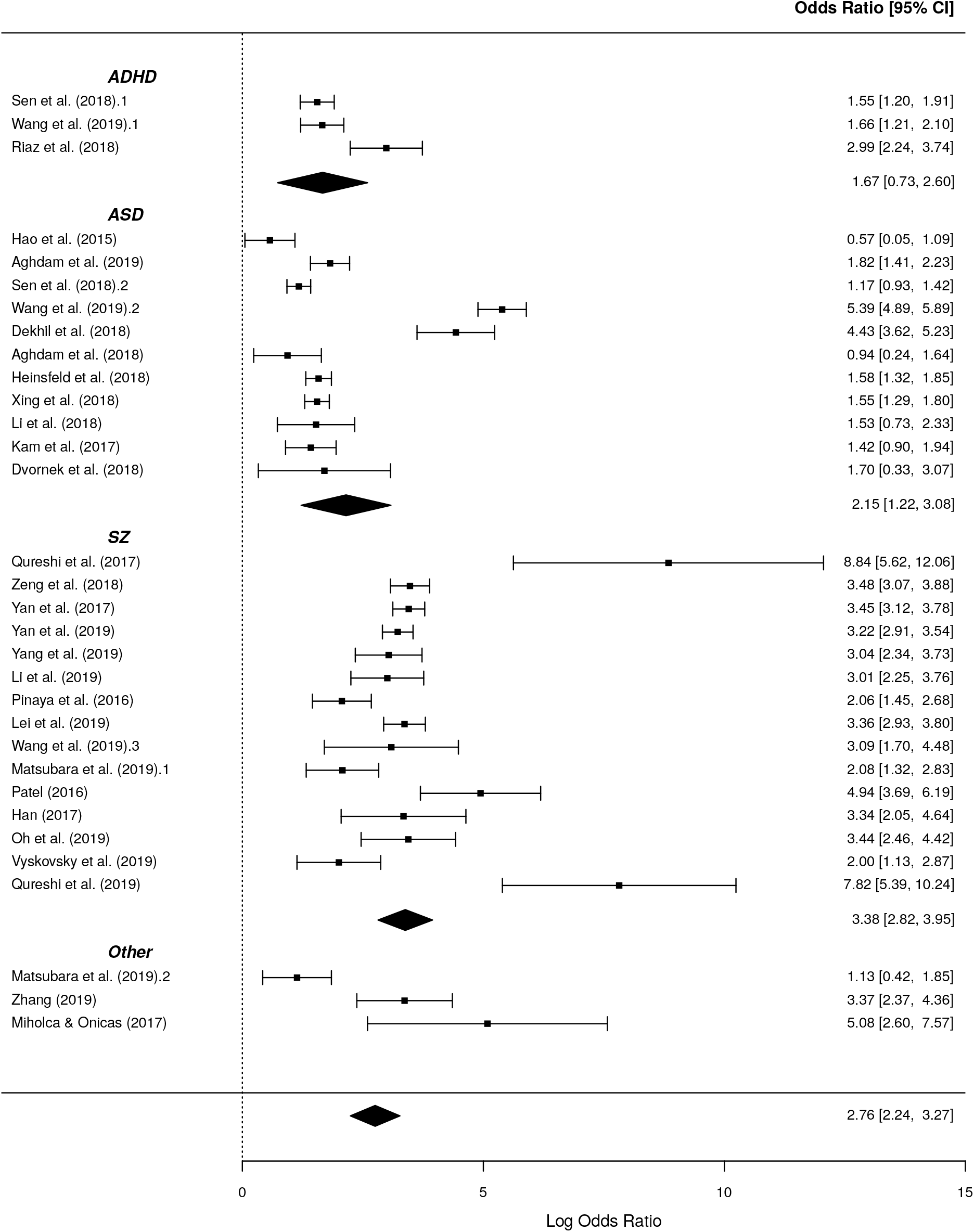
Univariate random-effect forest plots of log diagnostic odds ratio’s grouped per disorder.

## Discussion

### General conclusions from the existing literature

In the present review we systematically reviewed the literature applying deep learning methods to neuroimaging data for psychiatric disorders. Despite many promising results, the clinical use of DL to neuroimaging data to aid disease diagnosis for psychiatric disorders is still in its infancy. Given the complexity of the problem, starting from inherently uncertain diagnostic labels to heterogeneous scanning protocols and preprocessing, this is perhaps not surprising. Nevertheless, in recent years many studies have applied DL techniques to classify psychiatric disorders. While the body of literature on ASD, SZ or ADHD is increasing steadily, only a few studies have applied DL on other disorders such as MDD. It seems that the large, publicly available datasets are driving research as many of the included studies were based on ABIDE, COBRE and ADHD-200 datasets. Furthermore, the way that these datasets provide the neuroimaging data seems to influence what kind of features have been used as input for DL. For example, the ABIDE offers preprocessed timecourses for various atlas parcellations and many ASD studies use atlas extracted timecourses or FC matrices as input. In contrast, in SZ studies the input is highly heterogeneous. Even though multiple studies are using the same datasets, it remains difficult to compare performances and to identify optimal models or feature input. Various studies still use different subsets of the available dataset due to different quality checks or preferences. Furthermore, due to the rapid development of DL techniques and the wealth of preprocessing and parameter choices, there is a large heterogeneity in models used and features engineered.

Only a few studies have directly compared either differently engineered features or different modality approaches, making a definite conclusion on specific inputs difficult. Resting-state fMRI seems to be used most often, but whenever structural MRI is used, it seems to result in similar accuracies. The vast majority applies a form of feature engineering to the data, instead of developing end-to-end models for MRI that could learn features from the raw data.

From the three main disorders discussed, SZ seems to obtain the highest classification performance. There are several non-exclusive possibilities that may explain the differences in performance. One possibility is that the labels are more reliable. However, the inter rater reliability (IRR) for SZ appears lower than for ASD and ADHD^58^. Yet, it is important to note that also IRR of these different disorders are difficult to compare as they have been assessed in different settings.^58^ Another possibility is that the samples were more homogenous. Figure 5 indicates that the samples for SZ were smaller and obtained at fewer sites. This could have reduced the clinical heterogeneity within the patient group that is associated with higher accuracy^59^, as well as the heterogeneity of the imaging data. This is also consistent with the observation that accuracy was higher with smaller sample sizes (Figure 5), which is in line with reviews for standard ML.^60,61^ However, this pattern was absent for SZ and even positive for ADHD, suggesting that the overall negative association was primarily driven by the negative trend for ASD studies. Another explanation may be that differences in brain anatomy and function are more distinct from healthy controls. This is supported by data from large-scale neuroimaging consortia that have reported larger differences in brain anatomy for SZ compared with ADHD and ASD ^62^.

Remarkably, only half of the studies report sensitivity or specificity performance measures, whereas these are important for interpretation, especially when case/control groups have unequal sample sizes ^27^, and required for the present meta-analysis based on diagnostic odds ratios. Also, many studies do not compare their model with a benchmark ML model. This hinders a comprehensive comparison and a quantitative analysis of all included studies. In the following section we will evaluate more good and bad practices.

### Good and bad practices

In general, it can be concluded that there are still quite some studies not adhering to the ten simple rules of individual differences in neuroimaging as proposed by Scheinost et al. (2019)^27^. The first rules describe the need for an out-of-sample prediction as it generates more accurate and generalizable models. Predictive models in neuroimaging can be susceptible to overfitting, the tendency to mistakenly fit sample-specific noise as if it were signal ^63^, especially since the number of predictors is usually far greater than the number of observations^64^. Common practice to deal with the curse of dimensionality in neuroimaging is feature selection or engineering. This should be done carefully as training and test set need to stay independent. In our reported tables the highest reported accuracies are often from studies doing feature selection and we need to carefully interpret these results. Doing feature selection is not a bad practice, but it should be done inside a cross validation loop or on a different dataset^27^. At least for three studies^37–39^ feature selection is done on the whole sample, leading to model ‘peeking’ into the test data, which may lead to optimistic results. For several other studies it was unclear whether this procedure was done properly^40,41,65^.

According to rule #3, cross validation should be used to test a model’s generalizability. Preferably even, the model should be tested on a separate, external dataset as this provides most evidence of model generalization, but this is often not feasible. Still, several studies only report accuracies based on a single train/test split^37,44,52,53,66^, therefore reporting an overly optimistic outcome and complicating comparisons with other studies. As the best practice for model generalizability is to use an independently collected dataset as test set, it is good practice to report leave site out validation as each site is an independent dataset. However, although many studies have used multi-site data, only few report leave-site-out cross-validation.

Moreover, not only accuracy should be reported, as overall accuracy may not translate well to accuracy for individual classes ^67^. Studies should at least also report sensitivity and specificity. Finally, although a considerable number of studies already shares data and code, this should become more common practice to facilitate external validation and model comparisons.

### Deep learning vs machine learning

Although DL has unlocked unprecedented success in various domains, its superiority as an analytical tool for neuroimaging in psychiatry is yet to be demonstrated. The added benefit of DL is its ability to capture nonlinear, subtle patterns, but the question arises whether these nonlinearities are exploitable at the currently available sample sizes and examined scales. Here we tried to examine the difference in performance between DL and conventional shallow ML models. As can be seen in Figure 6, for thirty-two out of thirty-five studies (91%) directly comparing DL to ML, the performance of DL models was higher. When statistically comparing the two techniques on the sixteen studies that did report sensitivity and specificity, which is necessary for meta-analysis on odds ratios^22^, no significant difference was obtained. This could merely be the result of insufficient power, or because the random-effect meta-regression with ML/DL covariate assumes that the data arise from a randomized design. This is a conservative approach as the results truly are paired results; they are obtained by application of both techniques to the same dataset. Unfortunately, there is a lack of meta-analytical models that account for pairing of test results^22^ and we can therefore not apply a more appropriate and possibly more liberal approach. We assume that a paired analysis will show significant better performance of DL techniques as DL performed better in 91,43%% of the included studies that compare both methods, and we have seen that comparisons of ML-DL within one disorder does lead to significant differences.

It is important to note that there may be a publication bias towards higher performances for DL models given the increased interest in DL and our search for DL papers specifically^68^. It is, for instance, likely that many included studies optimized parameters for their DL model but did not optimize parameters for their comparative ML model. The difference with and without optimisation can be large: In a study of Yang et al. (2019)^69^ they deployed a grid search method to find the optimal parameters for SVM. They obtained a cross validated accuracy of 71.98% on the entire ABIDE I, whereas Heinsfeld et al. (2018)^33^ report an accuracy of 65% using SVM on the ABIDE I when comparing its performance with an MLP. It is therefore important to have standardized procedures for fair comparison between DL and ML models. Furthermore, when comparing models, studies should test whether the difference in performance is significant. Given the increased interest in DL, it seems that most published articles are devoted to the development of new DL methods, instead of a neutral comparison.

### The challenge of heterogeneous multisite datasets

It is known that the application of shallow ML methods to neuroimaging data leads to higher and more stable accuracies as sample size increases^70^. We expected to see this trend in DL studies as well, especially since deep learning is a data-hungry technique, but this was not observed. This could be partly explained by a larger heterogeneity of large datasets with multiple scanning sites. Multisite data analysis limit the sensitivity for detecting abnormalities due to inhomogeneities in scanning parameters, subject populations and research protocols^71^.

Studies have indeed shown a drop in accuracy when shifting from homogeneous single-site data to multi-site data using the same model^72^. Nevertheless, heterogeneous input data reflect reality and should eventually lead to models that generalize better. Future studies should investigate how to incorporate the domain knowledge of multisite datasets or develop domain adaptation models that learn how to reduce the discrepancy between different sources.

### Strengths and limitations

We will shortly discuss the strengths and limitations of this review and meta-analysis. First of all, given the high interest in DL and rapid increase of DL studies in neuroimaging, there was a need for a systematic overview of DL applications. Given the rigorous search in technical and biologically oriented databases, we included a large amount of studies in an attempt to give a comprehensive overview. One important limitations of this overview is the lack of an extensive quality assessment of studies as is proposed by the Cochrane handbook^22^. This may have led to inclusion of studies of less quality and biased results. However, this enabled us to identify good and bad practices within the field. Furthermore, for a good comparison between ML and DL studies, a thorough investigation on publication bias is needed to establish the reliability of this trend in favour of DL.

The most important limitation for the meta-analysis is that we could only include a small amount of studies for quantitative analysis as most studies did not report sensitivity or specificity performances. Whenever more studies can be included, this would aid the generalization of our conclusions. Finally, performing a paired meta-regression would aid in the comparison of DL-ML performances, but appropriate methods for doing so still need to be developed.

### Conclusions and future directions

Effective and accurate diagnosis of psychiatric disorders is important for initiation and choice of effective treatment. This review confirms that deep learning on neuroimaging is a promising tool for development of biological diagnostic models that could aid diagnosis. While still in its early stages, the application of DL in neuroimaging for psychiatric disorders has shown promising results and obtained better performance than conventional shallow machine learning techniques. Nevertheless, several improvements are needed before the full potential of DL in psychiatric neuroimaging can be achieved. The fifty-five studies included in this review show a wide variety of patient characteristics, type of feature engineering and applied DL techniques which raises problems of generalizability. Due to these heterogeneous approaches we could not identify optimal models or approaches for bivariate classifications.

When choosing a model and reporting its accuracy, future studies should be mindful of the questions of interest they want to answer. If the aim is to develop a new DL model to improve performance, then an extensive, neutral comparison to benchmarked ML models should be made that includes important performance measures for diagnostic classification (including sensitivity and specificity). Alternatively, the aim could be to apply DL to different kinds of input data, as it can learn features from higher dimensional data than conventional ML techniques. Yet, we have seen that many studies still use linear feature engineered inputs, suggesting that the DL models are not used to their full potential. In general, studies should report extensive performance comparisons and keep in mind the ten rules for predictive modelling of individual differences^27^ including proper validation.

Since we found that publicly available datasets drive research, we suggest that our recommendations are best implemented bottom-up, by introducing standardized datasets, with standardized preprocessing protocols. Ideally, all code for models using these datasets should be publicly available. Similarly, not only the performance results should be reported, but the full data of (in)correct classification of all subjects should be made available to make a proper comparison between models. This would also help to identify subject IDs that are always classified wrong, which could aid to identify noise in the diagnostic labels.

In conclusion, neuroimaging research in psychiatry using deep learning is still evolving to achieve better performance. While there are important challenges to overcome, our findings provide preliminary evidence supporting the promising role of DL in the future development of biological neuroimaging biomarkers for psychiatric disorders.

#### Box 1

**A short introduction to deep learning**

Deep learning is a group of machine leaning methods that tries to learn features from the data by a hierarchical structure of consecutive nonlinear transformations. In the present review, we define a deep learning model as follows: a model is a deep model when it included two or more stacked layers and therefore learns features through a hierarchical learning process. Although deep learning is a subgroup of machine learning, when we refer to machine learning in this review, we refer to shallow machine learning models (such as support vector machines).

The building blocks of deep learning methods are called artificial neurons. The simplest form of an artificial neuron is the single-layer perceptron as proposed by Rosenblatt^86^. The perceptron takes inputs *x* that are multiplied with connection weights *w*. The sum of all weighted inputs is then passed onto a nonlinear activation function such as tanh, sigmoid or rectified linear unit (ReLu). The main idea of the perceptron is to learn the values of the weights *w* in order to make a decision whether the neuron should fire or not.

By stacking several of these neurons, a multi-layer perceptron (MLP) is created. An MLP is organized in layers; an input layer, one or more hidden layer(s) and an output layer. In the input layer, the input data is where the data is entered into the model, the hidden layers learn increasingly abstract features and the output layer assigns a class using the learned features. The type of network determines how these artificial neurons are connected to other neurons. The simplest form of a deep network is the multilayer perceptron (MLP), which is fully connected, meaning that each neuron is connected to all neurons of the previous layer. Each connection is associated with a weight value, reflecting the strength and direction (positive or negative) between two neurons in the network.

During training, the network learns through a gradient descent-based algorithm, that aims to find the optimal weights that lead to a minimal error between predicted and true outputs. The idea behind training with gradient descent is as follows: as training data is fed through the network, the gradient of the loss function is computed with respect to every weight using the chain rule, and the weights are changed using gradient descent.

#### Box 2

**Deep learning architectures**

Besides MLPs, there exists a wide variety of deep learning architectures. We will shortly discuss the most common architectures in neuroimaging here. For a more elaborate overview of methods see Jo et al. (2019)^14^ and Vieira et al.(2017) ^10^

A. Deep belief network (DBN) Whereas MLPs only have feedforward connections, the DBN has undirected connections between some layers. These undirected layers are called Restricted Boltzmann Machines (RBM) and can be trained both supervised and unsupervised.
B. Convolutional neural network (CNN) CNNs are mostly used in image recognition. They work by learning ‘convolutions’ or ‘filters’ to detect features. By convolving images, it reduces the data into a form that is easier to process, without losing critical information.
C. Recurrent neural network (RNN) RNNs do not only contain feedforward connections, but also feedback connections. These feedback connections allow the retainment of information from previous inputs (akin to a form of memory) to affect the current output. The most effective RNNs are gated RNNs such as long short-term memory (LSTM) and networks based on the gated recurrent unit (GRU).
D. Auto Encoder (AE) AE is an unsupervised learning method that is used to encode the data in a smaller latent representation. They consist of an encoder and decoder part and are trained by making the output value approximate to its input value.

## Data Availability

n/a

## Footnotes

1* Although deep learning is a subtype of machine learning, we use the term machine learning (ML) for conventional shallow ML techniques (see Box 1)

## Notes

### Competing Interest Statement

The authors have declared no competing interest.

### Funding Statement

No external funding was received

## References

1. Sheffield, J. M. & Barch, D. M. Cognition and resting-state functional connectivity in schizophrenia. Neuroscience and Biobehavioral Reviews (2016). doi:10.1016/j.neubiorev.2015.12.007

2. Mulders, G. et al. E-learning improves knowledge and practical skills in haemophiliapatients on home treatment: a randomized controlled trial. HAEMOPHILIA 18, 693–698 (2012).

3. Kennedy, D. P. & Courchesne, E. The intrinsic functional organization of the brain is altered in autism. Neuroimage (2008). doi:10.1016/j.neuroimage.2007.10.052

4. Rubinov, M. & Sporns, O. Complex network measures of brain connectivity: Uses and interpretations. Neuroimage (2010). doi:10.1016/j.neuroimage.2009.10.003

5. Gong, G. et al. Mapping anatomical connectivity patterns of human cerebral cortex using in vivo diffusion tensor imaging tractography. Cereb. Cortex 19, 524–536 (2009).

6. Orrù, G., Pettersson-Yeo, W., Marquand, A. F., Sartori, G. & Mechelli, A. Using Support Vector Machine to identify imaging biomarkers of neurological and psychiatric disease: A critical review. Neuroscience and Biobehavioral Reviews (2012). doi:10.1016/j.neubiorev.2012.01.004

7. Arbabshirani, M. R., Plis, S., Sui, J. & Calhoun, V. D. Single subject prediction of brain disorders in neuroimaging: Promises and pitfalls. Neuroimage 145, 137–165 (2017).

8. Lu, D. & Weng, Q. A survey of image classification methods and techniques for improving classification performance. International Journal of Remote Sensing (2007). doi:10.1080/01431160600746456

9. Samper-González, J. et al. Reproducible evaluation of classification methods in Alzheimer’s disease: Framework and application to MRI and PET data. Neuroimage (2018). doi:10.1016/j.neuroimage.2018.08.042

10. Vieira, S., Pinaya, W. H. L. & Mechelli, A. Using deep learning to investigate the neuroimaging correlates of psychiatric and neurological disorders: Methods and applications. Neuroscience and Biobehavioral Reviews (2017). doi:10.1016/j.neubiorev.2017.01.002

11. Plis, S. M. et al. Deep learning for neuroimaging: a validation study. Front. Neurosci. 8, 229 (2014).

12. Durstewitz, D., Koppe, G. & Meyer-Lindenberg, A. Deep neural networks in psychiatry. Molecular Psychiatry (2019). doi:10.1038/s41380-019-0365-9

13. Bzdok, D. & Meyer-Lindenberg, A. Machine Learning for Precision Psychiatry: Opportunities and Challenges. Biological Psychiatry: Cognitive Neuroscience and Neuroimaging (2018). doi:10.1016/j.bpsc.2017.11.007

14. Jo, T., Nho, K. & Saykin, A. J. Deep Learning in Alzheimer’s Disease: Diagnostic Classification and Prognostic Prediction Using Neuroimaging Data. Front. Aging Neurosci. 11, 220 (2019).

15. Page, A., Turner, J. T., Mohsenin, T. & Oates, T. Comparing raw data and feature extraction for seizure detection with deep learning methods. in Proceedings of the 27th International Florida Artificial Intelligence Research Society Conference, FLAIRS 2014 (2014).

16. Sen, B., Borle, N. C., Greiner, R. & Brown, M. R. G. A general prediction model for the detection of ADHD and Autism using structural and functional MRI. PLoS One 13, e0194856– e0194856 (2018).

17. Pinaya, W. H. L., Mechelli, A. & Sato, J. R. Using deep autoencoders to identify abnormal brain structural patterns in neuropsychiatric disorders: A large-scale multi-sample study. Hum. Brain Mapp. 40, 944–954 (2019).

18. Matsubara, T., Tashiro, T. & Uehara, K. Deep Neural Generative Model of Functional MRI Images for Psychiatric Disorder Diagnosis. IEEE Trans. Biomed. Eng. 66, 2768–2779 (2019).

19. Wang, Z., Sun, Y., Shen, Q. & Cao, L. Dilated 3D Convolutional Neural Networks for Brain MRI Data Classification. IEEE Access 7, 134388–134398 (2019).

20. Gatsonis, C. & Paliwal, P. Meta-analysis of diagnostic and screening test accuracy evaluations: Methodologic primer. American Journal of Roentgenology (2006). doi:10.2214/AJR.06.0226

21. Reitsma, J. B. et al. Bivariate analysis of sensitivity and specificity produces informative summary measures in diagnostic reviews. J. Clin. Epidemiol. (2005). doi:10.1016/j.jclinepi.2005.02.022

22. Macaskill, P., Gatsonis, C., Deeks, J., Harbord, R. & Takwoingi, Y. Chapter 10: Analysing and Presenting Results. in Cochrane Handbook for Systematic Reviews of Diagnostic Test Accuracy Version 1.0 (2010).

23. Ismail, M. et al. A new deep-learning approach for early detection of shape variations in autism using structural mri. in 2017 IEEE International Conference on Image Processing (ICIP) 1057–1061 (2017). doi:10.1109/ICIP.2017.8296443

24. Li, X. et al. 2-Channel convolutional 3D deep neural network (2CC3D) for fMRI analysis: ASD classification and feature learning. in 2018 IEEE 15th International Symposium on Biomedical Imaging (ISBI 2018) 1252–1255 (2018). doi:10.1109/ISBI.2018.8363798

25. Mwangi, B., Tian, T. S. & Soares, J. C. A review of feature reduction techniques in Neuroimaging. Neuroinformatics (2014). doi:10.1007/s12021-013-9204-3

26. Varoquaux, G. et al. Assessing and tuning brain decoders: Cross-validation, caveats, and guidelines. Neuroimage (2017). doi:10.1016/j.neuroimage.2016.10.038

27. Scheinost, D. et al. Ten simple rules for predictive modeling of individual differences in neuroimaging. NeuroImage (2019). doi:10.1016/j.neuroimage.2019.02.057

28. Dvornek, N. C., Yang, D., Ventola, P. & Duncan, J. S. Learning Generalizable Recurrent Neural Networks from Small Task-fMRI Datasets. Med. Image Comput. Comput. Assist. Interv. 11072, 329–337 (2018).

29. Dvornek, N. C., Ventola, P. & Duncan, J. S. Combining phenotypic and resting-state fMRI data for autism classification with recurrent neural networks. in 2018 IEEE 15th International Symposium on Biomedical Imaging (ISBI 2018) 725–728 (2018). doi:10.1109/ISBI.2018.8363676

30. Dvornek, N. C., Ventola, P., Pelphrey, K. A. & Duncan, J. S. Identifying Autism from Resting-State fMRI Using Long Short-Term Memory Networks. Mach. Learn. Med. imaging. MLMI 10541, 362–370 (2017).

31. Dekhil, O. et al. Using resting state functional MRI to build a personalized autism diagnosis system. in 2018 IEEE 15th International Symposium on Biomedical Imaging (ISBI 2018) 1381–1385 (2018). doi:10.1109/ISBI.2018.8363829

32. Guo, X. et al. Diagnosing Autism Spectrum Disorder from Brain Resting-State Functional Connectivity Patterns Using a Deep Neural Network with a Novel Feature Selection Method. Front. Neurosci. 11, 460 (2017).

33. Heinsfeld, A. S., Franco, A. R., Craddock, R. C., Buchweitz, A. & Meneguzzi, F. Identification of autism spectrum disorder using deep learning and the ABIDE dataset. NeuroImage Clin. (2018). doi:10.1016/j.nicl.2017.08.017

34. Aghdam, M. A., Sharifi, A. & Pedram, M. M. Diagnosis of Autism Spectrum Disorders in Young Children Based on Resting-State Functional Magnetic Resonance Imaging Data Using Convolutional Neural Networks. J. Digit. Imaging (2019). doi:10.1007/s10278-019-00196-1

35. Mellema, C., Treacher, A., Nguyen, K. & Montillo, A. Multiple Deep Learning Architectures Achieve Superior Performance Diagnosing Autism Spectrum Disorder Using Features Previously Extracted From Structural And Functional Mri. in 2019 IEEE 16th International Symposium on Biomedical Imaging (ISBI 2019) 1891–1895 (2019). doi:10.1109/ISBI.2019.8759193

36. Khosla, M., Jamison, K., Kuceyeski, A. & Sabuncu, M. R. Ensemble learning with 3D convolutional neural networks for functional connectome-based prediction. Neuroimage 199, 651–662 (2019).

37. Li, G., Liu, M., Sun, Q., Shen, D. & Wang, L. Early Diagnosis of Autism Disease by Multi-channel CNNs. Mach. Learn. Med. imaging. MLMI 11046, 303–309 (2018).

38. Yan, W. et al. Discriminating schizophrenia from normal controls using resting state functional network connectivity: A deep neural network and layer-wise relevance propagation method. in 2017 IEEE 27th International Workshop on Machine Learning for Signal Processing (MLSP) 1–6 (2017). doi:10.1109/MLSP.2017.8168179

39. Yan, W. et al. Discriminating schizophrenia using recurrent neural network applied on time courses of multi-site FMRI data. EBioMedicine (2019). doi:10.1016/j.ebiom.2019.08.023

40. Qureshi, M. N. I., Oh, J. & Lee, B. 3D-CNN based discrimination of schizophrenia using resting-state fMRI. Artif. Intell. Med. 98, 10–17 (2019).

41. Qureshi, M. N. I., Oh, J., Cho, D., Jo, H. J. & Lee, B. Multimodal discrimination of schizophrenia using hybrid weighted feature concatenation of brain functional connectivity and anatomical features with an extreme learning machine. Front. Neuroinform. 11, 1–14 (2017).

42. Lei, D. et al. Detecting schizophrenia at the level of the individual: relative diagnostic value of whole-brain images, connectome-wide functional connectivity and graph-based metrics. Psychol. Med. 1–10 (2019). doi:10.1017/S0033291719001934

43. Oh, K. et al. Classification of schizophrenia and normal controls using 3D convolutional neural network and outcome visualization. Schizophr. Res. (2019). doi:10.1016/j.schres.2019.07.034

44. Srinivasagopalan, S., Barry, J., Gurupur, V. & Thankachan, S. A deep learning approach for diagnosing schizophrenic patients. J. Exp. Theor. Artif. Intell. 00, 1–14 (2019).

45. Patel, P., Aggarwal, P. & Gupta, A. Classification of schizophrenia versus normal subjects using deep learning. in ACM International Conference Proceeding Series (Association for Computing Machinery, 2016). doi:10.1145/3009977.3010050

46. Chyzhyk, D., Savio, A. & Graña, M. Computer aided diagnosis of schizophrenia on resting state fMRI data by ensembles of ELM. Neural Networks 68, 23–33 (2015).

47. Hao, A. J., He, B. L. & Yin, C. H. Discrimination of ADHD children based on Deep Bayesian Network. in 2015 IET International Conference on Biomedical Image and Signal Processing (ICBISP 2015) 1–6 (2015). doi:10.1049/cp.2015.0764

48. Deshpande, G., Wang, P., Rangaprakash, D. & Wilamowski, B. Fully Connected Cascade Artificial Neural Network Architecture for Attention Deficit Hyperactivity Disorder Classification From Functional Magnetic Resonance Imaging Data. IEEE Trans. Cybern. 45, 2668–2679 (2015).

49. Wang, T. & Kamata, S. Classification of Structural MRI Images in Adhd Using 3D Fractal Dimension Complexity Map. in 2019 IEEE International Conference on Image Processing (ICIP) 215–219 (2019). doi:10.1109/ICIP.2019.8802930

50. Zou, L., Zheng, J., Miao, C., Mckeown, M. J. & Wang, Z. J. 3D CNN Based Automatic Diagnosis of Attention Deficit Hyperactivity Disorder Using Functional and Structural MRI. IEEE Access 5, 23626–23636 (2017).

51. Riaz, A. et al. Deep fMRI: AN end-to-end deep network for classification of fMRI data. in 2018 IEEE 15th International Symposium on Biomedical Imaging (ISBI 2018) 1419–1422 (2018). doi:10.1109/ISBI.2018.8363838

52. Kuang, D., Guo, X., An, X., Zhao, Y. & He, L. LNBI 8590 - Discrimination of ADHD Based on fMRI Data with Deep Belief Network. LNBI 8590, (2014).

53. Kuang, D. & He, L. Classification on ADHD with Deep Learning. in 2014 International Conference on Cloud Computing and Big Data 27–32 (2014). doi:10.1109/CCBD.2014.42

54. Zhang, J. et al. Three dimensional convolutional neural network-based classification of conduct disorder with structural MRI. Brain Imaging Behav. (2019). doi:10.1007/s11682-019-00186-5

55. Pominova, M. et al. Voxelwise 3D Convolutional and Recurrent Neural Networks for Epilepsy and Depression Diagnostics from Structural and Functional MRI Data. in 2018 IEEE International Conference on Data Mining Workshops (ICDMW) 299–307 (2018). doi:10.1109/ICDMW.2018.00050

56. Miholca, D. & Onicaş, A. Detecting depression from fMRI using relational association rules and artificial neural networks. in 2017 13th IEEE International Conference on Intelligent Computer Communication and Processing (ICCP) 85–92 (2017). doi:10.1109/ICCP.2017.8116987

57. Vyskovsky, R., Schwarz, D. & Kasparek, T. Brain Morphometry Methods for Feature Extraction in Random Subspace Ensemble Neural Network Classification of First-Episode Schizophrenia. Neural Comput. 31, 897–918 (2019).

58. Regier, D. A. et al. DSM-5 field trials in the United States and Canada, part II: Test-retest reliability of selected categorical diagnoses. Am. J. Psychiatry (2013). doi:10.1176/appi.ajp.2012.12070999

59. Schnack, H. G. & Kahn, R. S. Detecting neuroimaging biomarkers for psychiatric disorders: Sample size matters. Front. Psychiatry (2016). doi:10.3389/fpsyt.2016.00050

60. Woo, C., Chang, L. J., Lindquist, M. A. & Wager, T. D. Building better biomarkers?: brain models in translational neuroimaging. 20, 365–377 (2017).

61. Wolfers, T., Buitelaar, J. K., Beckmann, C. F., Franke, B. & Marquand, A. F. pattern recognition for neuroimaging-based psychiatric diagnostics. Neurosci. Biobehav. Rev. (2015). doi:10.1016/j.neubiorev.2015.08.001

62. Thompson, P. M. et al. ENIGMA and global neuroscience: A decade of large-scale studies of the brain in health and disease across more than 40 countries. Translational Psychiatry (2020). doi:10.1038/s41398-020-0705-1

63. Yarkoni, T. & Westfall, J. Choosing Prediction Over Explanation in Psychology: Lessons From Machine Learning. Perspect. Psychol. Sci. (2017). doi:10.1177/1745691617693393

64. Whelan, R. & Garavan, H. When optimism hurts: Inflated predictions in psychiatric neuroimaging. Biol. Psychiatry (2014). doi:10.1016/j.biopsych.2013.05.014

65. Wang, C., Xiao, Z., Wang, B. & Wu, J. Identification of Autism Based on SVM-RFE and Stacked Sparse Auto-Encoder. IEEE Access 7, 118030–118036 (2019).

66. Li, G. et al. Application of deep canonically correlated sparse autoencoder for the classification of schizophrenia. Comput. Methods Programs Biomed. 183, 105073 (2019).

67. Baldi, P., Brunak, S., Chauvin, Y., Andersen, C. A. F. & Nielsen, H. Assessing the accuracy of prediction algorithms for classification: An overview. Bioinformatics (2000). doi:10.1093/bioinformatics/16.5.412

68. Boulesteix, A. L., Lauer, S. & Eugster, M. J. A. A Plea for Neutral Comparison Studies in Computational Sciences. PLoS One (2013). doi:10.1371/journal.pone.0061562

69. Yang, X., Islam, M. S. & Khaled, A. M. A. Functional connectivity magnetic resonance imaging classification of autism spectrum disorder using the multisite ABIDE dataset. in 2019 IEEE EMBS International Conference on Biomedical & Health Informatics (BHI) 1–4 (2019). doi:10.1109/BHI.2019.8834653

70. Nieuwenhuis, M. et al. Classification of schizophrenia patients and healthy controls from structural MRI scans in two large independent samples. Neuroimage (2012). doi:10.1016/j.neuroimage.2012.03.079

71. Nielsen, J. A. et al. Multisite functional connectivity MRI classification of autism: ABIDE results. Front. Hum. Neurosci. (2013). doi:10.3389/fnhum.2013.00599

72. Kam, T.-E., Suk, H.-I. & Lee, S.-W. Multiple functional networks modeling for autism spectrum disorder diagnosis. Hum. Brain Mapp. 38, 5804–5821 (2017).

73. Akhavan Aghdam, M., Sharifi, A. & Pedram, M. M. Combination of rs-fMRI and sMRI Data to Discriminate Autism Spectrum Disorders in Young Children Using Deep Belief Network. J. Digit. Imaging 31, 895–903 (2018).

74. Xing, X., Ji, J. & Yao, Y. Convolutional Neural Network with Element-wise Filters to Extract Hierarchical Topological Features for Brain Networks. in 2018 IEEE International Conference on Bioinformatics and Biomedicine (BIBM) 780–783 (2018). doi:10.1109/BIBM.2018.8621472

75. Ktena, S. I. et al. Metric learning with spectral graph convolutions on brain connectivity networks. Neuroimage 169, 431–442 (2018).

76. Li, H., Parikh, N. A. & He, L. A Novel Transfer Learning Approach to Enhance Deep Neural Network Classification of Brain Functional Connectomes. Front. Neurosci. 12, 491 (2018).

77. Parisot, S. et al. Disease Prediction using Graph Convolutional Networks: Application to Autism Spectrum Disorder and Alzheimer’s Disease. (2018). doi:10.1016/j.media.2018.06.001

78. Anirudh, R. & Thiagarajan, J. J. Bootstrapping Graph Convolutional Neural Networks for Autism Spectrum Disorder Classification. (2017). doi:arXiv:1704.07487v2

79. Dakka, J. et al. Learning Neural Markers of Schizophrenia Disorder Using Recurrent Neural Networks. (2017).

80. Pinaya, W. H. L. et al. Using deep belief network modelling to characterize differences in brain morphometry in schizophrenia. Sci. Rep. 6, 38897 (2016).

81. Ulloa, A., Plis, S., Erhardt, E. & Calhoun, V. Synthetic structural magnetic resonance image generator improves deep learning prediction of schizophrenia. in 2015 IEEE 25th International Workshop on Machine Learning for Signal Processing (MLSP) 1–6 (2015). doi:10.1109/MLSP.2015.7324379

82. Han, S., Huang, W., Zhang, Y., Zhao, J. & Chen, H. Recognition of early-onset schizophrenia using deep-learning method. Appl. Informatics 4, (2017).

83. Yang, B. et al. Schizophrenia Classification Using fMRI Data Based on a Multiple Feature Image Capsule Network Ensemble. IEEE Access 7, 109956–109968 (2019).

84. Zeng, L. L. et al. Multi-Site Diagnostic Classification of Schizophrenia Using Discriminant Deep Learning with Functional Connectivity MRI. EBioMedicine 30, 74–85 (2018).

85. Kim, J., Calhoun, V. D., Shim, E. & Lee, J.-H. Deep neural network with weight sparsity control and pre-training extracts hierarchical features and enhances classification performance: Evidence from whole-brain resting-state functional connectivity patterns of schizophrenia. Neuroimage 124, 127–146 (2016).

86. Rosenblatt, F. The perceptron: A probabilistic model for information storage and organization in the brain. Psychol. Rev. (1958). doi:10.1037/h0042519

87. Moher, D. et al. Preferred reporting items for systematic reviews and meta-analyses: The PRISMA statement. PLoS Medicine (2009). doi:10.1371/journal.pmed.1000097

